# A systems serology analysis of correlates of protection against cholera

**DOI:** 10.1101/2022.06.08.22276167

**Authors:** Kirsten E. Wiens, Anita S. Iyer, Taufiqur R. Bhuiyan, Lenette L. Lu, Deniz Cizmeci, Matthew J. Gorman, Dansu Yuan, Rachel L. Becker, Edward T. Ryan, Stephen B. Calderwood, Regina C. LaRocque, Fahima Chowdhury, Ashraful I. Khan, Myron M. Levine, Wilbur H. Chen, Richelle C. Charles, Andrew S. Azman, Firdausi Qadri, Galit Alter, Jason B. Harris

## Abstract

Vibriocidal antibodies are the best characterized correlate of protection against cholera and are used to gauge immunogenicity in vaccine trials. However, there is no vibriocidal titer threshold associated with absolute protection against infection with *Vibrio cholerae* or with symptomatic disease in infected individuals. While other circulating antibody responses have also been associated with a decreased risk of *V. cholerae* infection, there has been no comprehensive comparison of correlates of protection against cholera. To address this, we analyzed 58 serum antibody biomarkers as correlates of protection against both *V. cholerae* infection and against cholera diarrhea in infected individuals. The study was performed in two cohorts: (1) household contacts of patients with cholera in Bangladesh, and (2) North American volunteers who were vaccinated with a single dose of CVD 103-HgR live oral cholera vaccine and then challenged with virulent *V. cholerae* O1 El Tor Inaba. In household contacts, we identified 20 antibody markers that were correlated with protection against *V. cholerae* infection, though there was overlap between distributions. Conditional random forest models identified serum antibody-dependent complement deposition, targeting the *V. cholerae* O1 antigen, as the most predictive individual correlate of protection from infection, while vibriocidal antibody titers were less predictive. The model that most accurately predicted protection from infection included five biomarkers, with a cross-validated area under the curve (cvAUC) of 79% (95% CI 73-85). Similarly, in North American volunteers who were challenged with *V. cholerae* after vaccination, a different five-biomarker model predicted protection from the development of cholera diarrhea with a cvAUC of 78% (95% CI 66-91). Thus, while several new biomarkers predict protection better than vibriocidal titers, it remains difficult to consistently predict whether an individual will be protected from future mucosal infection or symptoms using current serologic markers.

## Introduction

Circulating antibodies that correlate with protection from infection or disease are valuable for understanding whether segments of the population are likely to be protected due to prior infection or vaccination. For cholera, the most well characterized and widely used correlate of protection is the serum vibriocidal antibody titer (Iyer and Harris, 2021). Vibriocidal titers remain elevated in the blood for up to a year or longer following infection (Azman et al., 2019), but are not thought to represent a mechanistic form of mucosal immunity (Iyer and Harris, 2021). Serum IgA antibodies specific to several *Vibrio cholerae* antigens are also correlated with protection from infection (Harris et al., 2008; Kaisar et al., 2021). While intestinal IgA antibodies could represent a direct form of protection, circulating IgA responses are short-lived following *V. cholerae* infection (Azman et al., 2019). For all of these known correlates, there is no titer that is associated with absolute protection.

In addition to serum vibriocidal and IgA responses, extensive household-based studies conducted in Dhaka, Bangladesh have shown that age, ABO blood group, and certain species in the intestinal microbiota are associated with infection and disease severity (Harris et al., 2008; Levade et al., 2021; Midani et al., 2018; Ritter et al., 2019; Weil et al., 2009). However, taken together, these variables do not entirely account for the variation in susceptibility or disease severity seen in this population. This residual variation is likely attributable to a combination of individual-level risk factors, exposure histories, and unmeasured or undetected immune responses. The degree to which unmeasured immune responses contribute to variation in susceptibility is unknown, and no comprehensive comparison of the multiple known correlates of protection against cholera has been conducted.

Here, our objective was to perform a comprehensive analysis of antibody-mediated correlates of protection from both *V. cholerae* infection and cholera diarrhea. Specifically, we asked: (1) Can we identify novel serum correlates of susceptibility to infection and/or cholera diarrhea after exposure to *V. cholerae* following both prior infection and vaccination? and (2) Do pre-infection biomarkers allow for accurate identification of who will be infected after exposure and/or who will go on to develop symptoms?

To this end, we conducted a systems serology analysis (Arnold and Chung, 2018; Chung and Alter, 2017) of 58 pre-infection serum biomarkers in two cohorts of individuals that were followed prospectively to monitor *V. cholerae* infection and cholera diarrhea. The biomarkers evaluated included all previously identified antigens associated with protective immunity as well as functional markers of antibody-mediated antibacterial immunity. The first cohort included contacts in the household of patients who had cholera in Dhaka, Bangladesh, where cholera is endemic (Levade et al., 2021; Weil et al., 2009). The second cohort consisted of North American volunteers who were vaccinated with a live attenuated oral cholera vaccine and subsequently challenged with *V. cholerae* (Chen et al., 2016). We tested single- and multi-biomarker panels in prediction using several machine-learning approaches. Our work identifies several novel biomarkers, as well as both the utility and limitations of using serum biomarkers to predict susceptibility to mucosal infection.

## Results

### Household contacts of index cholera cases

We included samples and data from 261 contacts of *V. cholerae* O1-Ogawa-infected cholera cases in 180 households in Dhaka, Bangladesh. The characteristics of this cohort are shown in Table 1. The cohort included 4 individuals who were under five years old, all of whom went on to develop infection. 77 of the 261 contacts had a positive stool culture (i.e., became infected) after exposure to a case in the household. Among the infected contacts, 26 were symptomatic and 51 were asymptomatic.

**Table 1.** Characteristics of the household contacts of index cholera cases in Dhaka, Bangladesh.

|  |  | Total |  | Infected |  |
| --- | --- | --- | --- | --- | --- |
| Category | Group | N | % | N | % |
| Sex | Female | 134 | 51.3 | 40 | 29.9 |
|  | Male | 127 | 48.7 | 37 | 29.1 |
| Age group | 0-4 | 4 | 1.5 | 4 | 100.0 |
|  | 5-17 | 74 | 28.4 | 25 | 33.8 |
|  | 18-49 | 164 | 62.8 | 45 | 27.4 |
|  | 50-71 | 19 | 7.3 | 3 | 15.8 |
| Self-reported history of prior cholera | No | 251 | 96.2 | 76 | 30.3 |
|  | Yes | 10 | 3.8 | 1 | 10.0 |
| Vibriocidal titer $\geq 320$ upon enrollment | No | 226 | 86.6 | 74 | 32.7 |
|  | Yes | 35 | 13.4 | 3 | 8.6 |
| Index cholera case received antibiotics | No | 181 | 69.3 | 60 | 33.1 |
|  | Yes | 80 | 30.7 | 17 | 21.2 |
| Relationship to index cholera case | Child | 48 | 18.4 | 18 | 37.5 |
|  | Sibling | 47 | 18.0 | 15 | 31.9 |
|  | Spouse | 61 | 23.4 | 15 | 24.6 |
|  | Parent | 101 | 38.7 | 29 | 28.7 |
| Diarrhea duration in index case | 0-6 days | 67 | 25.7 | 17 | 25.4 |
|  | 7-20 days | 92 | 35.2 | 23 | 25.0 |
|  | 14-20 days | 38 | 14.6 | 12 | 31.6 |
|  | 21-73 days | 64 | 24.5 | 25 | 39.1 |

### Correlates of protection in household contacts

We examined 58 serum antibody biomarkers in household contacts upon enrollment. These included 54 antigen-isotype specific responses measured using a multiplex bead assay: IgA, IgA1, IgA2, IgM, IgG, IgG1, IgG2, IgG3, and IgG4 specific for *V. cholerae* O1 Ogawa OSP:BSA, O1 Inaba OSP:BSA, toxin coregulated pilus (TcpA), cholera holotoxin (CT-HT), cholera toxin B subunit (CtxB), and *V. cholerae* sialidase. These antigens represent dominant targets of the humoral immune response following cholera (Kauffman et al., 2016) (Azman et al., 2019). We also examined previous measurements of vibriocidal titers (Ritter et al., 2019) as well as three previously uninvestigated measures of functional antibody responses against cholera. These included measures of antibody-dependent complement deposition (ADCD), antibody-dependent cellular phagocytosis (ADCP), and antibody-dependent neutrophil phagocytosis (ADNP) targeting the *V. cholerae* O1 antigen. We verified that patients recovering from cholera mounted ADCD, ADCP and ADNP responses (Figure 1).

**Figure 1.**
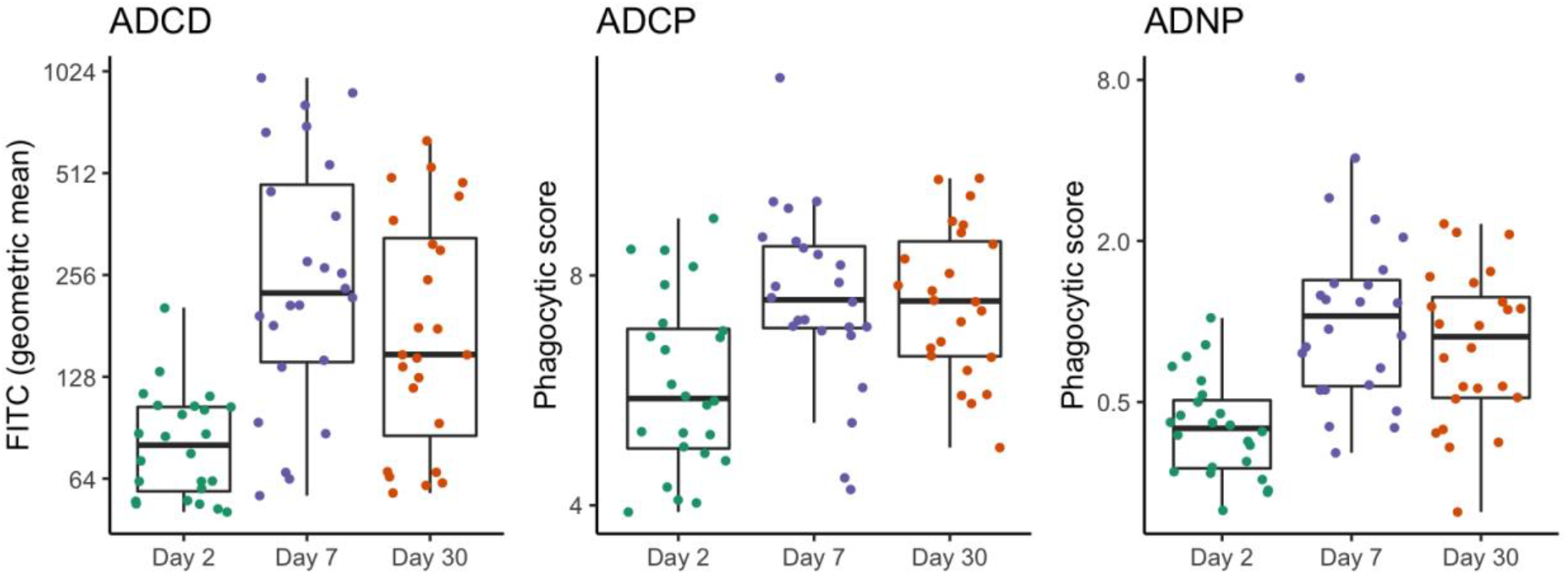
Dynamics of functional antibody responses targeting the *V. cholerae* O1 antigen following *V. cholerae* infection. Antibody-dependent complement deposition (ADCD), cellular phagocytosis (ADCP), and neutrophil phagocytosis (ADNP) in serum from 24 index cholera cases upon enrollment (i.e., Day 2 post presumed symptom onset) and days 7 and 30 post onset. Each point represents the geometric mean of complement deposition and phagocytic scores, respectively, across three replicates. Boxplots show 50% (median), 25%, and 75% quartiles; whiskers represent approximate 95% confidence intervals for comparing medians (McGill et al., 1978).

In single biomarker analyses, after adjusting for age and household clustering, 20 biomarkers were elevated among household contacts that remained uninfected (Figure S1) and hence were correlates of protection from subsequent infection. Among individual biomarkers, the risk of becoming infected with *V. cholerae* O1 was particularly low in individuals with elevated ADCD, TcpA IgG2, and TcpA IgA2 titers (Figure 2). Correlates of protection from symptoms among infected individuals were distinct from correlates of protection from initial infection (Figure S2), though sample sizes were smaller (n=77) and there was substantial overlap in antibody distributions between outcomes (Figure S1).

**Figure 2.**
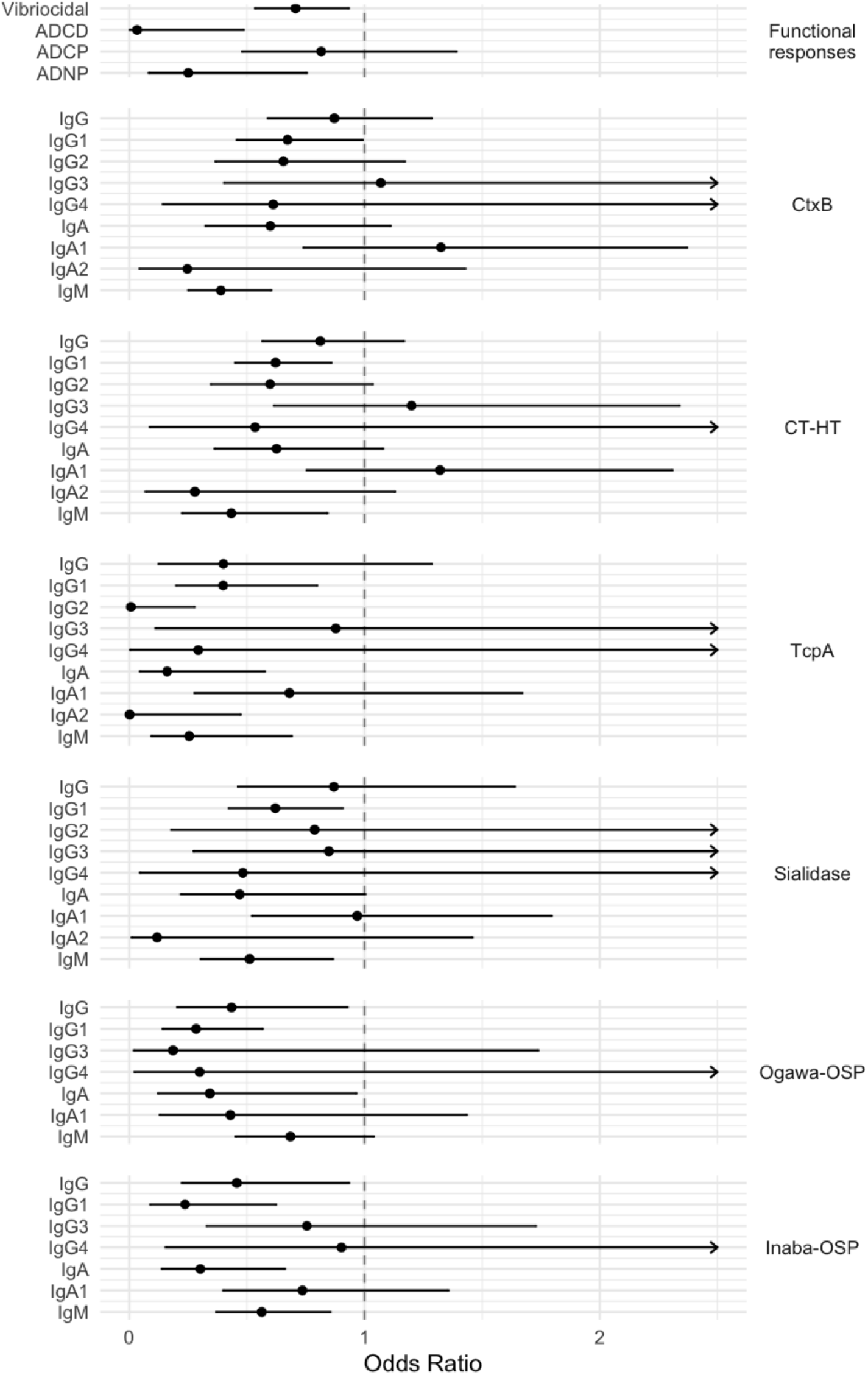
Risk of infection in household contacts of an index cholera case. Odds ratio of becoming infected among household contacts of an index cholera case for every 2-fold increase in baseline (Day 2) antibody titers, after adjusting for age and household clustering. Mean and 95% confidence interval are shown for each biomarker analyzed independently. Arrows indicate where upper confidence interval extends beyond the graph. Isotype is indicated on the left and antigen on the right for binding antibody titers. Biomarkers for which logistic model fitted values were very close to 1 are excluded.

In an unsupervised multivariate analysis, we found limited separation of infection outcomes using all antibody responses (Figure S3), which were generally highly correlated (Figure S4). We then used conditional random forest models, designed to handle a large number of highly correlated variables (Strobl et al., 2007), to examine which biomarkers were most important for classifying individuals by outcome. ADCD was ranked most important for classifying infections, and there were several features with higher scores than vibriocidal titers (Figure 3).

**Figure 3.**
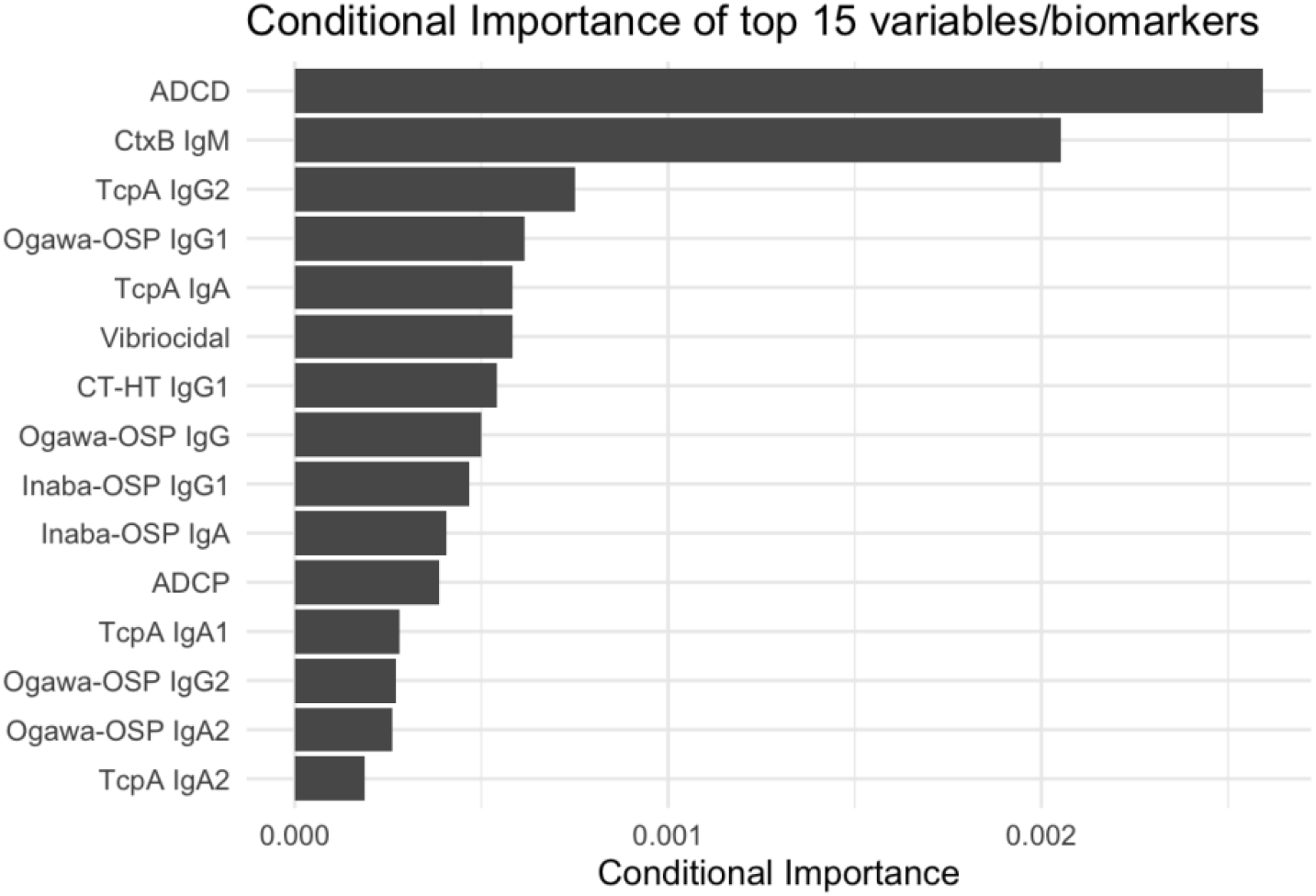
Biomarkers important for classifying household contacts by infection outcome. Top 15 biomarkers important in classifying household contacts of index cholera cases as infected (i.e., becoming stool culture positive) vs. uninfected (i.e., remaining stool culture negative). Biomarkers are ranked by importance scores calculated using conditional random forest classification models.

To test the validity of the classification models, we performed leave-one-out cross-validation for models constructed using different subsets of biomarkers and age. The model that most accurately predicted infection included five of the most informative biomarkers, with a cross-validated area under the curve (cvAUC) of 79% (95% CI 73-85) (Figure 4). This corresponded to a positive predictive value (PPV) of 0.85 and a negative predictive value (NPV) of 0.53. A model including ADCD and age performed comparably (cvAUC 74% (95% CI 68-81)) with PPV 0.83 and NPV 0.52. The vibriocidal model was less predictive (cvAUC 54% (95% CI 47-61)) with PPV 0.74 and NPV 0.26 (Figure 4). These results were consistent when we performed classification using an ensemble or “super learner” of three different classification models (random forest, penalized logistic regression, and support vector machine) (Figure S5).

**Figure 4.**
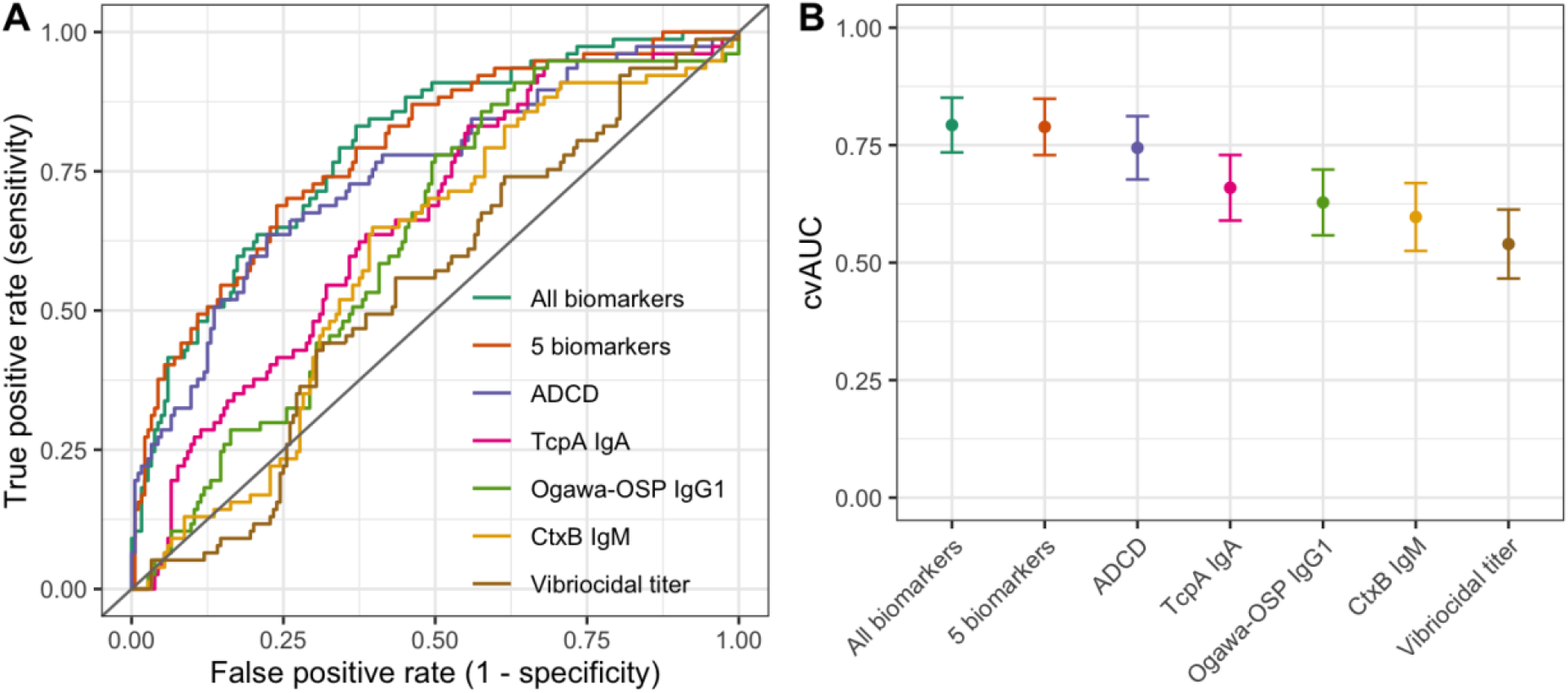
Predicting infection outcome among household contacts using different subsets of biomarkers. **A)** Cross-validated receiver operator curves (cvROC) for classifying household contacts of index cholera cases that remain uninfected vs. become infected using random forest models with different subsets of biomarkers and age. “5 biomarkers” corresponds to five of the top biomarkers selected via conditional importance, including ADCD, CtxB IgM, TcpA IgG2, Ogawa-OSP IgG1, and Sialidase IgG1. True and false positive rates calculated using leave-one-out cross-validation. **B)** Cross-validated area under the curve (cvAUC) corresponding to the models in A. Influence-curve based 95% confidence intervals are shown for the cvAUC estimates.

### Correlates of protection in challenged vaccine recipients

While household contacts of cholera cases have high risk of exposure in a real-world setting, a limitation of this study design for examining protection is that the degree and duration of exposure are unknown and varied among the study participants. To examine whether the primary correlates of protection for household contacts were consistent in vaccinees and in the setting of a controlled exposure, we replicated the analysis in a cohort of North American volunteers that received a live attenuated oral cholera vaccine (CVD 103-HgR) and were then challenged with approximately 1 × 10^5^ colony forming units of the wild type *V. cholerae O1* El Tor Inaba strain N16961 (Chen et al., 2016). This cohort included 34 individuals challenged 10 days post vaccination and 33 individuals challenged 90 days post vaccination (Table 2). Participants were 18-45 years old, 16 (24%) were female, and 20 (30%) developed mild to severe diarrhea following exposure (Table 2).

**Table 2.** Characteristics of North American volunteers who were vaccinated and then challenged with *V. cholerae*.

| Characteristic | Category | N | No qualifying diarrhea | Mild | Moderate | Severe |
| --- | --- | --- | --- | --- | --- | --- |
| Challenge day post vaccination | 10 | 34 | 29 (85.3%) | 3 (8.8%) | 1 (2.9%) | 1 (2.9%) |
|  | 90 | 33 | 18 (54.5%) | 11 (33.3%) | 2 (6.1%) | 2 (6.1%) |
| Age group | 18-25 | 15 | 12 (80%) | 3 (20%) | 0 (0%) | 0 (0%) |
|  | 26-35 | 34 | 24 (70.6%) | 7 (20.6%) | 1 (2.9%) | 2 (5.9%) |
|  | 36-45 | 18 | 11 (61.1%) | 4 (22.2%) | 2 (11.1%) | 1 (5.6%) |
| Sex | Female | 16 | 11 (68.8%) | 3 (18.8%) | 0 (0%) | 2 (12.5%) |
|  | Male | 51 | 36 (70.6%) | 11 (21.6%) | 3 (5.9%) | 1 (2%) |

We measured serum antibody responses for the same set of biomarkers as above, apart from ADCP (which was not measured due to limited sample volumes). As with the household contacts, we found no separation of vaccinees by diarrhea outcome in an unsupervised analysis of day-of-challenge serum biomarkers (Figure S6) and titers were highly correlated (Figure S7). In contrast to the household contacts, CT-HT IgA and CtxB IgA responses were ranked most conditionally important for classifying by diarrhea status (Figure 5). As with the household contacts, several features ranked higher than vibriocidal titers (Figure 3, Figure 5). Differences in biomarker importance between the cohorts persisted when we restricted the household contacts to the same age groups as the vaccinees (Figure S8). Relative ranking of biomarkers differed when we subset the vaccinees to those challenged on either day 10 or day 90 (Figure S9), though sample sizes were small in the subsets (Table 2) and uncertainty was high (Figure S10).

**Figure 5.**
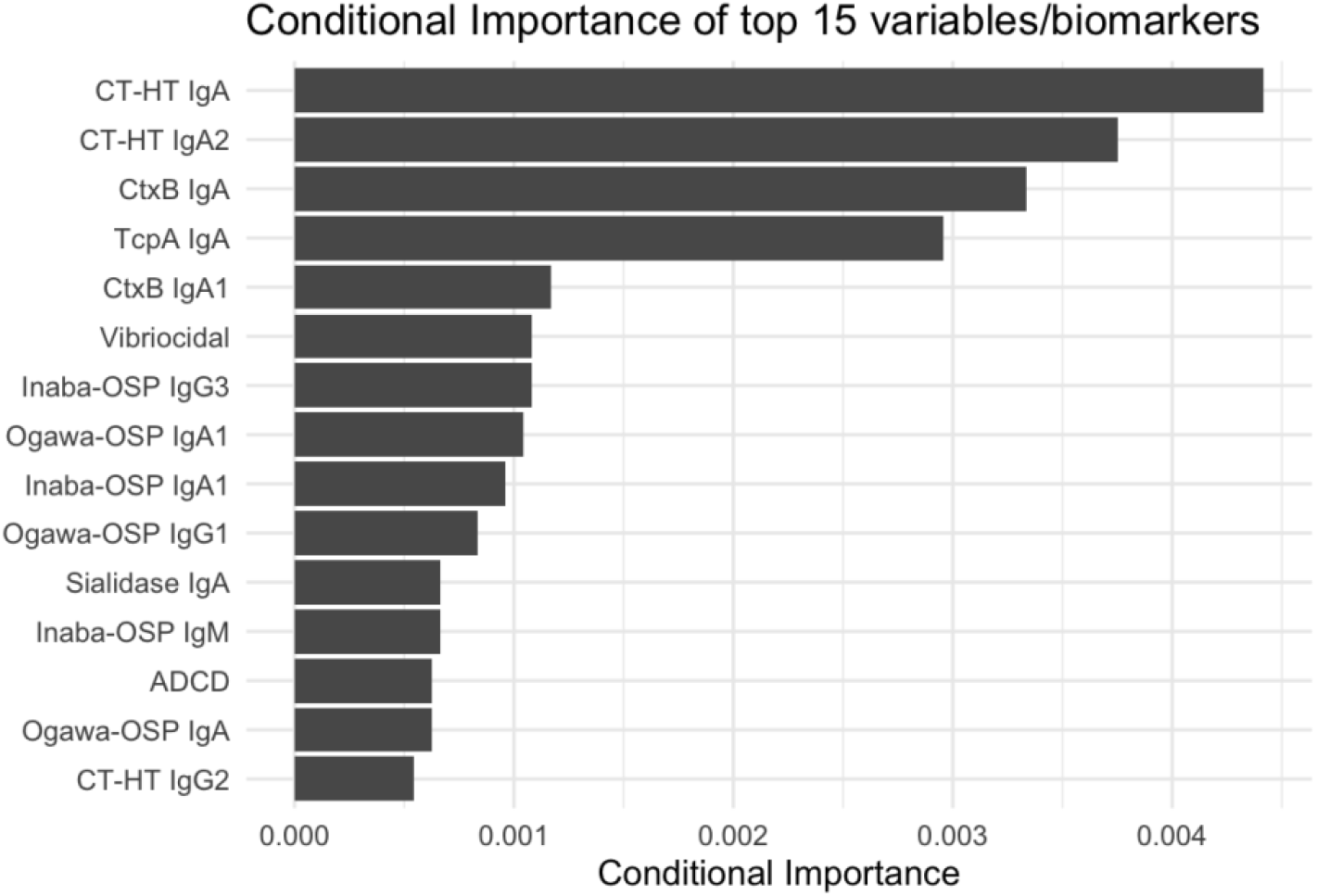
Biomarkers important for classifying vaccinees by whether they developed diarrhea following *V. cholerae* challenge. Top 15 biomarkers important in classifying vaccinees by developing either no qualifying diarrhea or mild to severe diarrhea following *V. cholerae* challenge. Biomarkers were selected using serum collected from participants on the day of challenge. Biomarkers are ranked by importance scores calculated using conditional random forest classification models.

We found comparable cross-validated results for the vaccinees as the household contacts. The model that most accurately predicted developing cholera diarrhea included five of the top biomarkers and age with cvAUC 78% (95% CI, 66-91%) with a PPV of 0.83 and NPV of 0.65 (Figure 6). Models that included only age and either CT-HT IgA2 or CtxB IgA performed comparably, followed by vibriocidal titers and then ADCD and TcpA IgA. The fold-increase in vibriocidal titers by day 10 following vaccination was less predictive of cholera diarrhea than day-of-challenge titers (Figure S11). These results were consistent when classification was performed using the ensemble model (Figure S12). Furthermore, the 5-biomarker fitted with the household contact data (Figure 4) performed nearly as well when predicting cholera diarrhea in the vaccinees with cvAUC 77% (95% CI, 64-90%) (Figure S13). In contrast, the 5-biomarker model fitted with the vaccinee data (Figure 6) performed poorly when predicting *V. cholerae* infection in the household contacts with cvAUC 60% (95% CI, 52-67%) (Figure S13).

**Figure 6.**
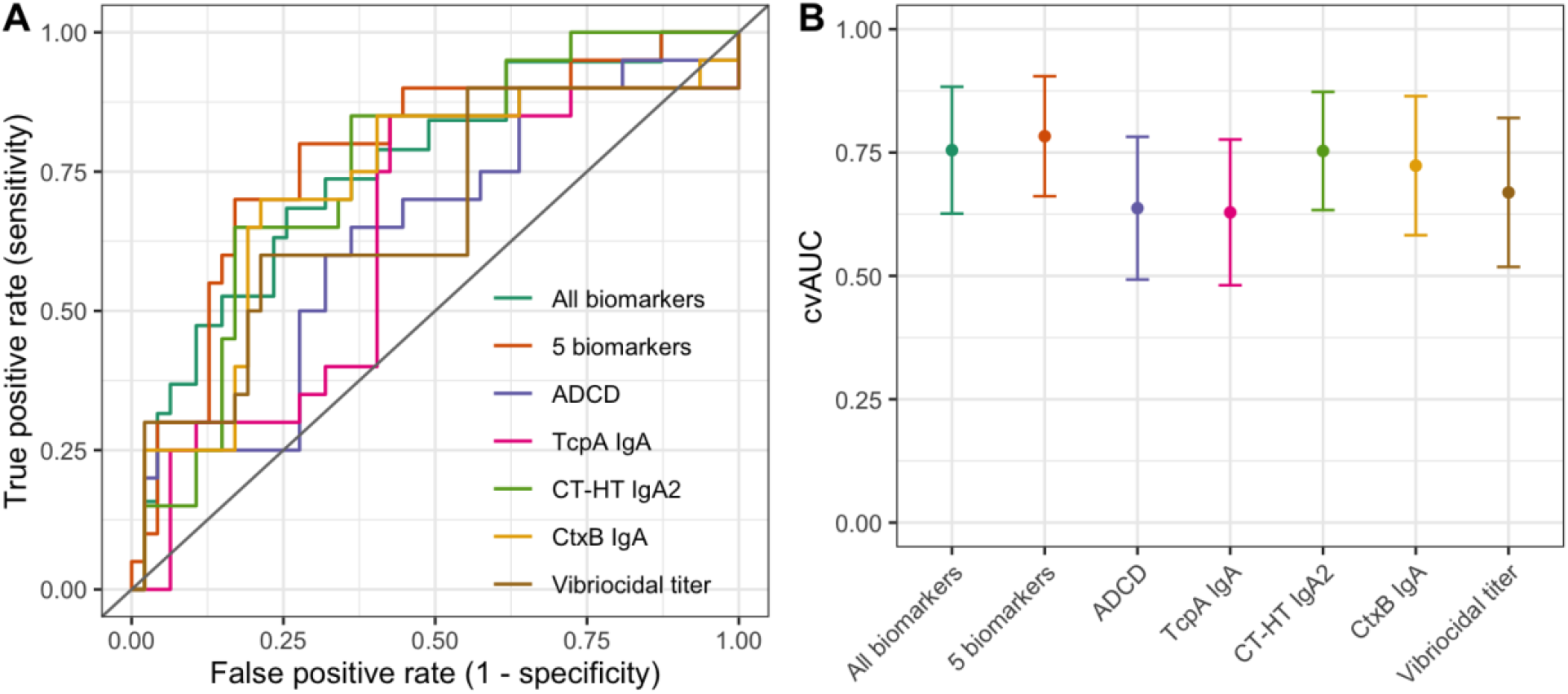
Predicting whether vaccinees develop diarrhea using different subsets of biomarkers. **A)** Cross-validated receiver operator curves (cvROC) for classifying vaccinees that develop diarrhea vs. those that do not using random forest models with different subsets of biomarkers and age. “5 biomarkers” corresponds to five of the top biomarkers selected via conditional importance, including CT-HT IgA, CtxB IgA, CT-HT IgA2, TcpA IgA, and Sialidase IgA2. True and false positive rates calculated using leave-one-out cross-validation. **B)** Cross-validated area under the curve (cvAUC) corresponding to the models in A. Influence-curve based 95% confidence intervals are shown for the cvAUC estimates.

## Discussion

Vibriocidal antibody titers are considered the best accepted correlate of protection against *V. cholerae* and are used as a proxy for predicting vaccine efficacy in Phase I/II clinical trials for cholera vaccines (Iyer and Harris, 2021). Using a systems serology approach, we identified 19 additional biomarkers that were also associated with reduced risk of infection in household contacts of patients with cholera. In models evaluating multiple immune measures, vibriocidal antibodies were less important in classifying risk of infection than other markers. This was true both in a cohort of household contacts and in a cohort of challenged cholera vaccinees. Our finding, that numerous markers individually and in combination are more predictive of protection from infection than vibriocidal titers, challenges the current standard that vibriocidal antibodies are indeed the best available correlate of protection against cholera.

Though we identified many antibody correlates of protection against cholera, our ability to distinguish between immunologically protected and susceptible individuals using current serologic markers was not perfect. This may reflect non-immunologic factors that influence the outcome of infection, or the inherent challenges involved in using serum biomarkers to predict mucosal immunity. Some of the unexplained protection may be attributable to *V. cholerae* O1-specific memory B cell responses, which have been shown to be associated with protection even in the absence of elevated levels of circulating antibodies (Haney et al., 2018; Patel et al., 2012). In addition, secretory IgA and innate immune signaling at the mucosal surface play important roles in modulating susceptibility (Karlsson et al., 2013; Weil et al., 2019) but cannot be measured directly in serum. Systems serology approaches to distinguishing active vs. latent tuberculosis (Lu et al., 2016) and mild vs. severe SARS-CoV-2 (Bartsch et al., 2021) were more predictive than ours, with cross-validated accuracies of up to 91-97% for certain outcomes. In contrast, a systems serology analysis of protection from typhoid fever found predictive accuracies similar to ours (73-86%) (Jin et al., 2020). These discrepancies could reflect inherent differences in immunity in the lungs compared to the gut, or differences between intracellular and extracellular pathogens, and how those responses correlate with circulating antibodies.

Despite lack of a perfect predictive model, we were able to identify robust correlates of protection in cholera. Some of these correlates had been identified previously (Iyer and Harris, 2021) while others, such as ADCD, are novel. Interestingly, ADCD was the single most predictive single marker of protection in household contacts and shares several similarities the vibriocidal titer marker. Both ADCD and vibriocidal titers are complement-dependent measures of functional antibody-mediated immunity. However, while vibriocidal titers reflect the downstream bactericidal activity of antibodies which activate complement via the classical pathway, ADCD is a bead-based, antigen-specific measure of C3 binding to *V. cholerae* O1-targeted antibodies. While there is ongoing debate whether circulating antibodies, transuded from the vascular compartment, could mediate complement dependent killing of *V. cholerae* and other enteric pathogens on the surface of the intestinal mucosa (Iyer and Harris, 2021), it is notable that these predictive markers both implicate complement-dependent antibody responses. Considering this, and that exposure of *V. cholerae* to O-antigen binding IgG induces bacterial responses which protect against complement mediated killing (Baranova et al., 2018), raises the possibility that complement could play a role in protective immunity against *V. cholerae*.

In this study, we did not find a ‘universal signature’ or single set of biomarkers that was optimal for classifying outcomes across both cohorts (household contacts and challenged vaccinees) or outcomes (infection and illness). However, the model which best predicted susceptibility in household contacts also predicted susceptibility in vaccinees with similar accuracy. In contrast, a model trained to predict protection in challenged vaccinees in a 10 to 90-day window did not accurately predict susceptibility among household contacts. This may reflect that a model trained in a single experimental setting is less likely to identify broadly applicable correlates of protection than models based on observed conditions in a cholera-endemic population. In other words, the household contact model may reflect correlates of protection which are applicable across a wider range of circumstances (e.g., different times between exposures, different strains of the bacteria, different populations) and to reflect multiple immune pathways which may lead to protection against cholera.

This study has limitations. We do not know if uninfected household contacts of index cholera cases were exposed to sufficient *V. cholerae* through contaminated water or food sources to be considered exposed yet protected. This may contribute to the variation seen in this cohort and may explain why predictive accuracy was slightly higher for vaccinees with known exposure status. In addition, we relied on previous measurements of vibriocidal titers in this cohort. In a subset of samples where we were able to repeat the vibriocidal assays, we found that the old and new responses were highly correlated (Figure S14) and gave comparable results in prediction (Figure S15). This suggests that changes over time in sample quality or handling may not have affected the other biomarkers but does not eliminate the possibility. Lastly, the conditional permutation method we used to rank biomarkers in the random forest models has some inherent instability and may give preference to highly correlated variables (Debeer and Strobl, 2020). However, the consistency of the results from this approach with that of ensemble of three models (Figure S5, Figure S12) supports the robustness of our overall conclusions.

In summary, we have identified a set of serum biomarkers that can be used to monitor susceptibility to *V. cholerae* infection and symptoms, with varying sensitivity and specificity. These new markers could allow for flexibility in designing studies of population susceptibility to infection, provided that predictive accuracy metrics are used to adjust final estimates. In addition, future studies could examine whether antibody markers in alternative samples such as stool and/or saliva correlate with mucosal immunity and allow for more accurate prediction. Finally, these findings illustrate the need to design better observational studies that allow us to more accurately measure *V. cholerae* exposure, for example by including questionnaires on frequency and duration of interactions with index cases in addition to measures of infectiousness in household surveys (Coit et al., 2019). Such analyses would provide important insights into how immunity and other risk factors contribute to both individual outcomes and secondary transmission.

## Materials and Methods

### Study populations and sample collection

#### Case-ascertainment cohorts in Dhaka, Bangladesh

We included samples and data from participants in Dhaka, Bangladesh from Dec. 2006 to Sept. 2018 (Ritter et al., 2019). Symptom histories, blood, and fecal samples were collected prospectively from household contacts of hospital-ascertained cholera cases in Dhaka, Bangladesh. Households were defined as individuals who shared the same cooking pot for ≥3 days. Stool samples and rectal swabs were tested for presence of *V. cholerae*. Blood specimens were tested for serum antibody titers and ABO blood group. For this study, we included 261 household contacts who were stool negative upon enrollment, did not report diarrhea symptoms in the preceding week or at enrollment, and had sufficient sample material for laboratory analyses. We excluded infected contacts whose *V. cholerae* serotype did not match the index case and contacts with “possible infections” (i.e., stool negative with ≥4-fold increase in vibriocidal titer and/or diarrhea symptoms).

#### Human challenge trial in North American volunteers

We included samples and data from 67 North American volunteers with no history of cholera who were vaccinated with 5 × 10^8^ colony-forming units (CFU) of the live attenuated oral cholera vaccine CVD 103-HgR and then challenged with approximately 5 × 10^5^ *V. cholerae* O1 El Tor Inaba strain N16961 on either 10 (n = 34) or 90 (n = 33) days post vaccination (Chen et al., 2016). Challenged participants were monitored for diarrhea symptoms and each stool was graded; all loose stools (grades 3-5) were weighed, and volumes calculated.

### Outcomes and definitions

#### Case-ascertainment cohorts in Dhaka, Bangladesh

The primary outcome in the household contacts cohort was *V. cholera* infection. Infection was defined as developing a positive stool culture result on day 7 or 30 after enrollment of the household’s index cholera case. A secondary outcome among infected household contacts was developing symptoms. Symptoms included watery diarrhea (≥3 loose or watery stools in a 24-hr period), abdominal pain, vomiting, and/or fever.

#### Human challenge trial in North American volunteers

The primary outcome in the vaccine challenge cohort was developing symptomatic cholera diarrhea. Diarrhea was defined as ≥2 loose stools of ≥200 mL each or a single loose stool of ≥300 mL over a 48-hour period. Moderate diarrhea was passage of ≥3 L and severe diarrhea was passage of ≥5 L loose stool (Chen et al., 2016).

### Laboratory methods

#### Antigens

*V. cholerae* Inaba and Ogawa OSP:BSA conjugates, TcpA, and *V. cholerae* sialidase were derived as previously described (Charles et al., 2017). *V. cholerae* holotoxin (List Biological, catalog 100B) and CtxB (Sigma, catalog C9903) were purchased from commercial vendors as indicated.

#### Luminex assay for antigen-specific antibody responses

We measured antigen-specific Ig class (IgG, IgA, IgM) and subclass (IgG1, IgG2, IgG, IgG4, IgA1, IgA2) responses against antigens using a customized Luminex assay. Each antigen was conjugated to unique, phycoerythrin (PE)-labeled, MagPlex-C Microsphere (Luminex Corp, Austin, TX) bead region at a concentration of 5 µg antigen per 1 × 10^6^ microspheres using the principles of carbodiimide coupling and as per manufacturer’s instructions (Luminex, xMAP® Antibody Coupling kit, catalog 40-50016).

Plasma samples were heat-inactivated by incubation at 56°C for 30 minutes and diluted in an assay buffer (0.1% BSA in 1X DPBS) to a final dilution of 1:100. Five µl of diluted plasma and 45 µl of diluted antigen-conjugated beads were added to a black 384-well polystyrene plate (Greiner Bio-One™, Monroe, NC) at a final concentration of 15 beads per µl (total: 675 beads per well). Plates were incubated overnight at 4°C with shaking (800 rpm) and then sonicated and washed thrice with 0.1%BSA in 1X PBS, 0.05% Tween20 before addition of PE-conjugated anti-human IgG, IgM and IgA (Southern Biotech; 40 µl per well) for a final dilution of 1:154. After incubation at 1 hour with shaking, plates were sonicated and washed thrice with 0.1% BSA in 1X PBS, 0.05% Tween20. The beads were resuspended in 40µl of sheath fluid (Fisher Scientific, Waltham, MA) and analyzed using Bioplex/Flexmap machines (BioRad). Samples were tested in duplicate, and data were analyzed as geometric mean fluorescence intensities.

#### Antigen biotinylation

Ogawa OSP:BSA antigen (2 mg/ml stock) and Inaba OSP:BSA (2.5mg/ml) was biotinylated at 20mM excess as per manufacturer’s instructions (Thermofisher Scientific, catalog: A39256) followed by removal of unbound biotin using Zeba Spin column 7K MWCO (Thermofisher, catalog: 89882).

#### Antibody-dependent neutrophil phagocytosis (ADNP assay)

Biotinylated Ogawa OSP:BSA antigen (2 mg/ml stock) and Inaba OSP:BSA (2.5mg/ml) was conjugated with 1.0µm yellow/green fluorescent neutravidin labeled microspheres (Thermofisher, catalog F8776) at 1:1 ratio and incubated at 37°C for 2 hours. The antigen-conjugated beads were pelleted, washed twice, and resuspended in 0.1% BSA in 1X DPBS and used within 4 days. Ten µl of heat inactivated (56°C for 30 mins) plasma diluted 1:4 in 0.1% BSA in 1X DPBS was added per well to a 96-well plate (Costar catalog:3799) followed by the addition of 10 µl/well of antigen-bead conjugate. Plates were incubated at 37°C 5% CO_2_ for 2 hours.

Blood collected from healthy donors was lysed using ammonium-chloride-potassium lysis buffer (150 mM NH_4_Cl, 10 mM KHCO3, 0.1 mM Na_2_EDTA, pH 7.4) at 1:10 ratio and incubated for 5 mins at room temperature. Cells were pelleted at 500×g for 5 mins, washed with cold 1X DPBS and resuspended in RPMI with 10% Fetal Calf Serum and 1X Penicillin/Streptomycin. Granulocytes were resuspended at a concentration of 2.5 × 10^5^ cells/ml. At the end of the 2-hour incubation, beads were pelleted, washed twice with 0.1% BSA in 1X DPBS and supernatants flicked. 200ul of resuspended cells (50,000 total cells) were added per well to the 96-well plate and plates were incubated at 37°C for 4 hours. Cells were pelleted (5 min, 4°C, 500 ×g), stained with CD66b-PacBlue (Biolegend), washed, and fixed with 4% paraformaldehyde. Samples were acquired within 48 hours on BD LSR Fortessa.

Neutrophil bead internalization was quantified using FlowJo (FlowJo, LLC) software by gating for granulocytes (FSC/SSC)/neutrophils (CD66b+)/bead+neutrophils (FITC+). Phagocytic score was calculated by multiplying the percentage of neutrophils that internalized the beads with geometric mean fluorescence intensity of bead+ neutrophils+ cells divided by 10,000. Samples were run in singlicate and results are an average of 3 independent experiments run using 3 different blood donors.

#### Antibody-dependent complement deposition (ADCD assay)

12 µl of biotinylated Ogawa OSP:BSA or Inaba OSP:BSA antigen was conjugated with 1.0 µm red fluorescent, neutravidin labeled microspheres (Thermofisher, catalog F8775) per 96-well plate at 37°C for 2 hours. The antigen:conjugated beads were pelleted and washed twice with 0.1%BSA in 1X DPBS followed by resuspension in 1200ul of 0.1% BSA in 1X DPBS. They were used within 4 days.

Ten µl of heat inactivated (56°C for 30 mins) plasma diluted 1:4 in 5% BSA in 1X DPBS was added to a 96-well plate (Costar catalog:3799) followed by the addition of 10 µl of antigen-bead conjugate and incubation at 37°C, 5% CO_2_. Four µl of reconstituted guinea pig complement (Cedarlane; catalog CL4051) diluted 1:50 in Gelatin Veronal Buffer was added per well to the 96-well plate and incubated at 37°C for 20 mins. At the end of incubation, beads were pelleted at 2000×g for 10 mins and then washed twice with cold 15mM EDTA in 1X DPBS.

Fifty µl of FITC labeled goat anti-guinea pig C3 diluted 1:200 in 1X PBS was added to the plate and incubated in the dark at room temperature for 15 mins. Plates were washed twice, and the beads were resuspended in 100µl PBS. Geometric MFI of FITC gated on red, neutravidin beads were calculated using FlowJo software. Samples were tested in singlicate and results are an average of 3 independent experiments.

#### Antibody-dependent cellular phagocytosis

Ogawa antigen bead conjugates were prepared as described for the ADNP assay. THP-1 cells were maintained in RPMI 1640 media (ATCC) containing 2 mM L-Glutamine (Corning), 10% Fetal Bovine Serum (Sigma), 10 mM HEPES (Corning), 55 μM beta-mercaptoethanol (Gibco), and 1X Penicillin/Streptomycin (Corning). A volume of 10 μL diluted human monoclonal antibody or plasma sample was added to each well, and immune complexes were formed over a 2-hour incubation at 37°C. After washing to remove non-specific unbound antibody, THP-1 cells were added (200 μL/well) at a concentration of 1.25 × 10^5^ cells/mL (2.5 × 10^4^ cells/well) and incubated with the immune complexed beads for 16 hours at 37°C. Cells were then fixed with 4% PFA and acquired on a BD LSRFortessa flow cytometer. The phagocytic score was calculated by multiplying the percentage of bead-positive cells by the geometric mean fluorescence intensities of the bead-positive cells divided by 10,000.

### Analytical methods

#### Single-marker correlates of infection

All analyses were performed in the R Statistical Software v4.0.3 (R Core Team, 2020). We examined risk of infection (i.e., developing a positive stool culture result) among all household contacts after exposure to an index cholera case, and risk of developing symptoms among infected household contacts. Odds ratios of developing infection or symptomatic cholera for each 2-fold increase in baseline biomarker titers were calculated using generalized estimating equations using the gee-package (Carey et al., 2019), adjusting for household clustering and age. Biomarkers were centered and scaled prior to analysis. Biomarkers in logistic models with fitted values very close to 1 were excluded.

#### Unsupervised analysis

We examined whether participants were separated by individual outcomes (*V. cholerae* infection and cholera diarrhea) using generalized principal component analysis (GLM-PCA), which allows for dimension reduction in non-normally distributed data. Analyses were run using the glmpca-package with a negative binomial likelihood (Townes et al., 2019).

#### Supervised analyses

We used conditional random forest models to examine which baseline biomarkers were most important for classifying individuals that went on to develop infection vs. those that remained uninfected and, of those infected, which went on to develop symptoms vs. which remained asymptomatic in the household contacts. We used the same approach to classify cholera diarrhea vs. no qualifying diarrhea in the vaccine challenge cohort. Models were run with 1000 trees and class weights to account for imbalances in the data, and otherwise default settings from the party package were used (Strobl et al., 2008, 2007). Conditional permutation importance was used to rank variables via the perimp-package (Debeer and Strobl, 2020).

We used leave-one-out cross-validation to examine the predictive validity of the conditional random forest models for the outcomes described above using different subsets of biomarkers and age. Predictions were visualized using receiver operator curves (cvROC) built using the pROC-package (Robin et al., 2011). The area under those curves (cvAUC) and corresponding influence curve-based confidence intervals were calculated using the cvAUC-package (LeDell et al., 2014). In addition, we compared cvROC and cvAUC for the conditional random forest models to that of an ensemble of three different machine learning models: random forest, penalized logistic regression, and support vector machine. Analyses were run using class weights to account for imbalances in the dataset and otherwise default settings in the SuperLearner-package (Polley et al., 2021).

## Data availability

All data generated in this study are available in the manuscript and Supplementary file 1. All input datasets and analytical code are available at: https://github.com/HopkinsIDD/cholera-systems-serology and https://doi.org/10.5281/zenodo.6626079.

## Data Availability

All data produced in the present work are contained in the manuscript and are available online at https://github.com/HopkinsIDD/cholera-systems-serology

## Acknowledgements

This research was supported by the icddr,b and extramural grants from the National Institutes of Health, including the National Institute of Allergy and Infectious Diseases, (R01 AI137164 [J.B.H., R.C.C., G.A., F.Q.], R01 AI106878 [E.T.R., F.Q.], R01 AI130378 [T.R.B.], R01 AI135115 [A.S.A, K.E.W.]). icddr,b is thankful to the donors for their support to its research efforts. icddr,b is also grateful to the governments of Bangladesh, Canada, Sweden, and the United Kingdom for providing core/unrestricted support.

## Competing interests

The authors declare that they have no competing interests.

## Ethics

This study was approved by icddr,b Ethical Review Committee and Massachusetts General Hospital’s Institutional Review Boards.

## Notes

### Competing Interest Statement

The authors have declared no competing interest.

### Author Declarations

This study was approved by International Centre for Diarrhoeal Disease Research Bangladesh Ethical Review Committee and Massachusetts General Hospital Institutional Review Boards

