## Supplementary file 1 for "A systems serology analysis of correlates of protection against cholera"

Includes Figures S1-S15

**Figure S1. Antibody titers upon enrollment among household contacts of index cholera cases**

Functional and antigen-isotype-specific binding antibody responses among household contacts in the case-ascertainment studies that remained uninfected (green), that became infected but didn't develop symptoms (purple), and that became infected and developed symptoms (red). Each point represents the geometric mean of antibody titers across 2-3 replicates for each contact upon enrollment (Day 2). Boxplots show 50% (median), 25%, and 75% quantiles; whiskers represent approximate 95% confidence intervals for comparing medians (McGill et al. (1978)). The * indicates differences between infected and uninfected were significant after adjusting for age and household clustering (also see Figure 2). The ^ indicates differences between infected contacts that developed symptoms and that remained asymptomatic were significant after adjusting for age and household clustering (also see Figure S2).

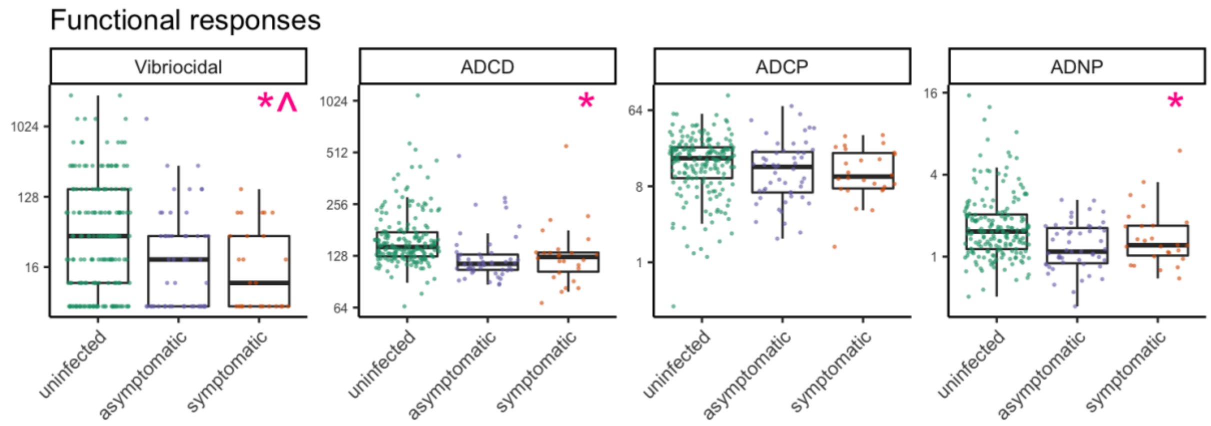

CtxB

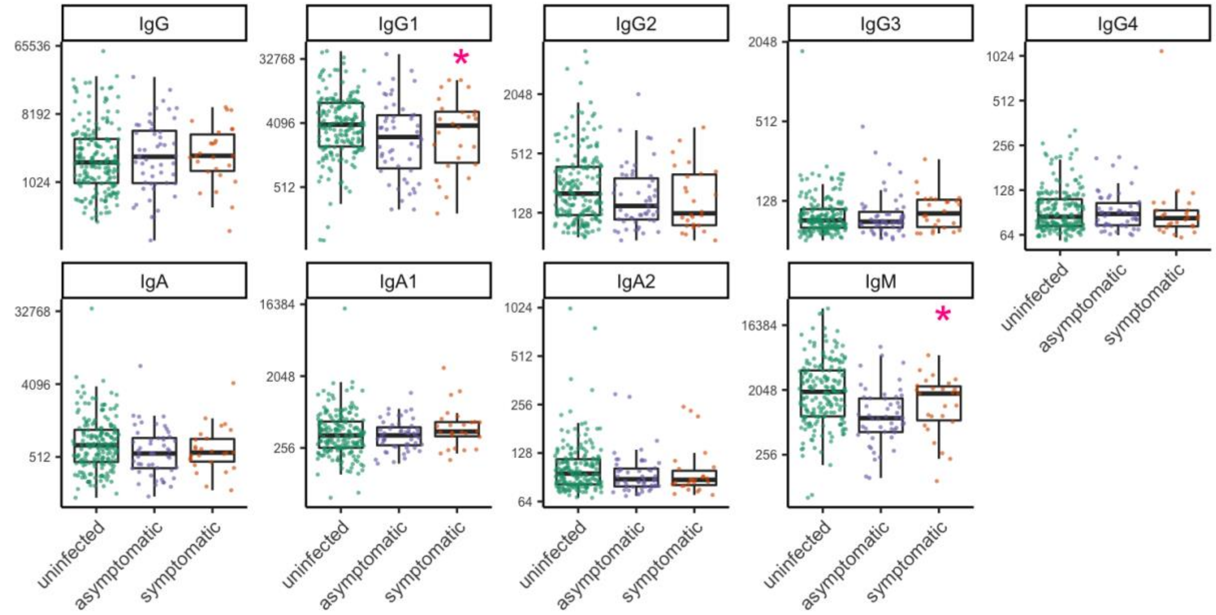
IgG
IgG1
IgG2
IgG3
IgG4
IgA
IgA1
IgA2
IgM
65536
8192
1024
32768
4096
512
2048
128
256
64
16384
uninfected
asymptomatic
symptomatic

CT-HT

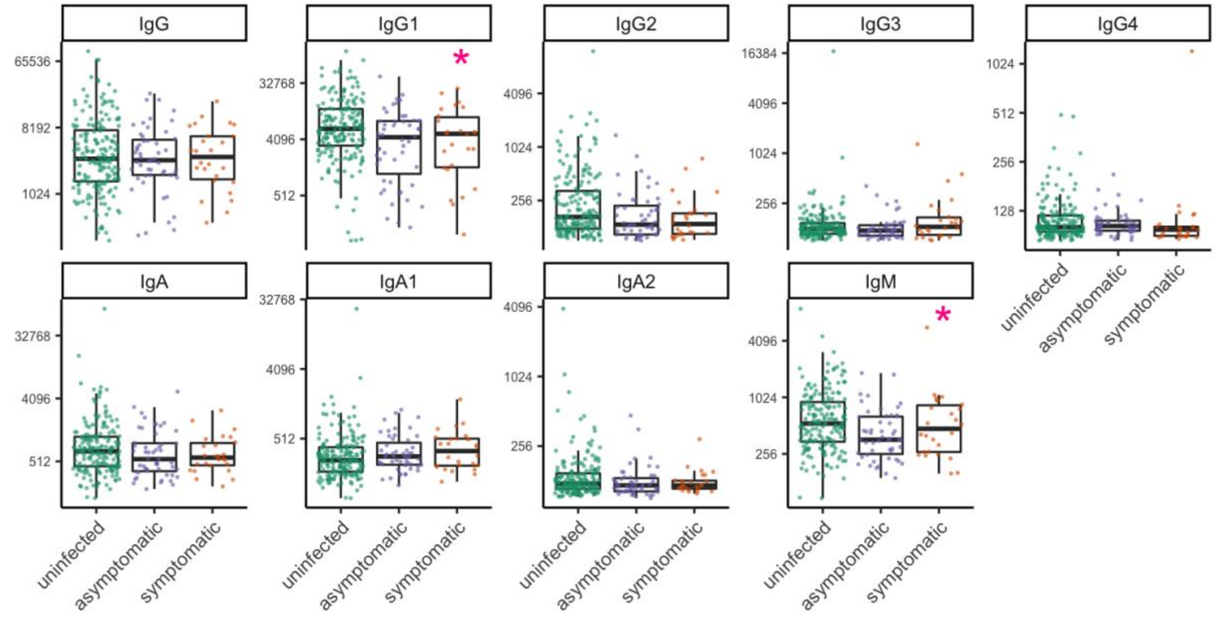
IgG
IgG1
IgG2
IgG3
IgG4
IgA
IgA1
IgA2
IgM
65536
8192
1024
32768
4096
512
256
16384
128
uninfected
asymptomatic
symptomatic

TcpA

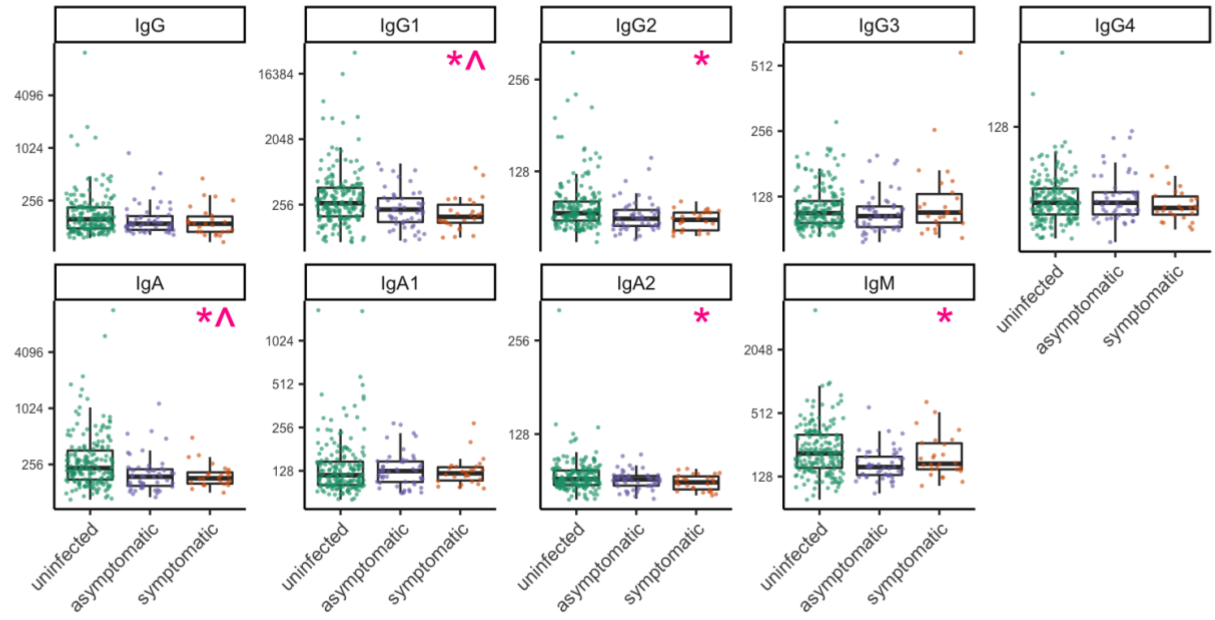


Sialidase

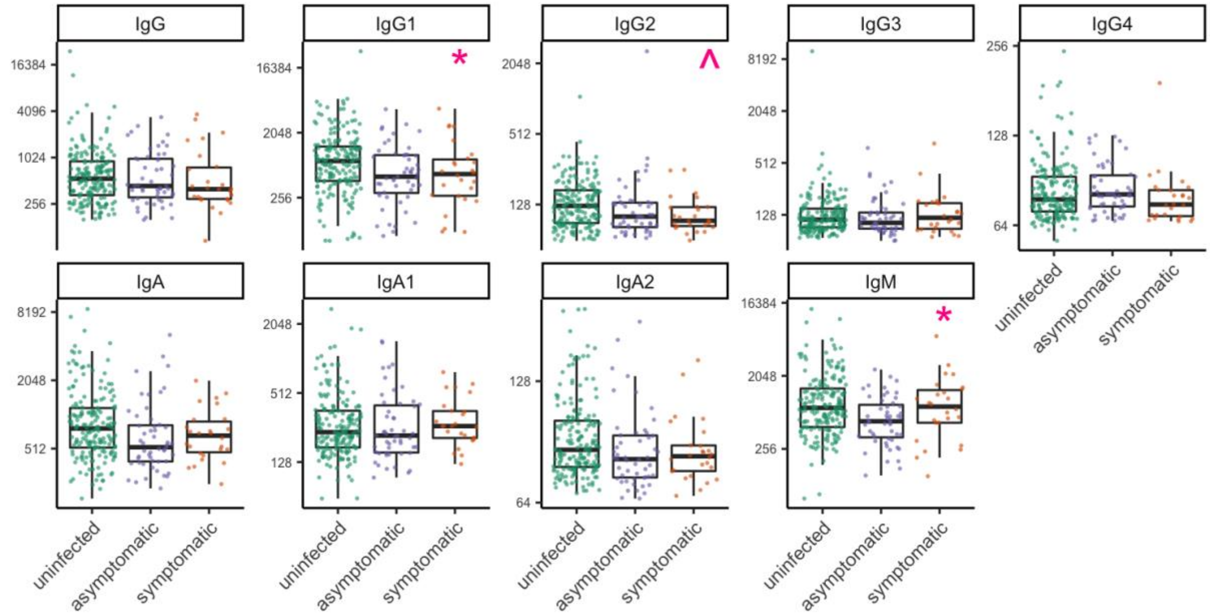

Ogawa-OSP

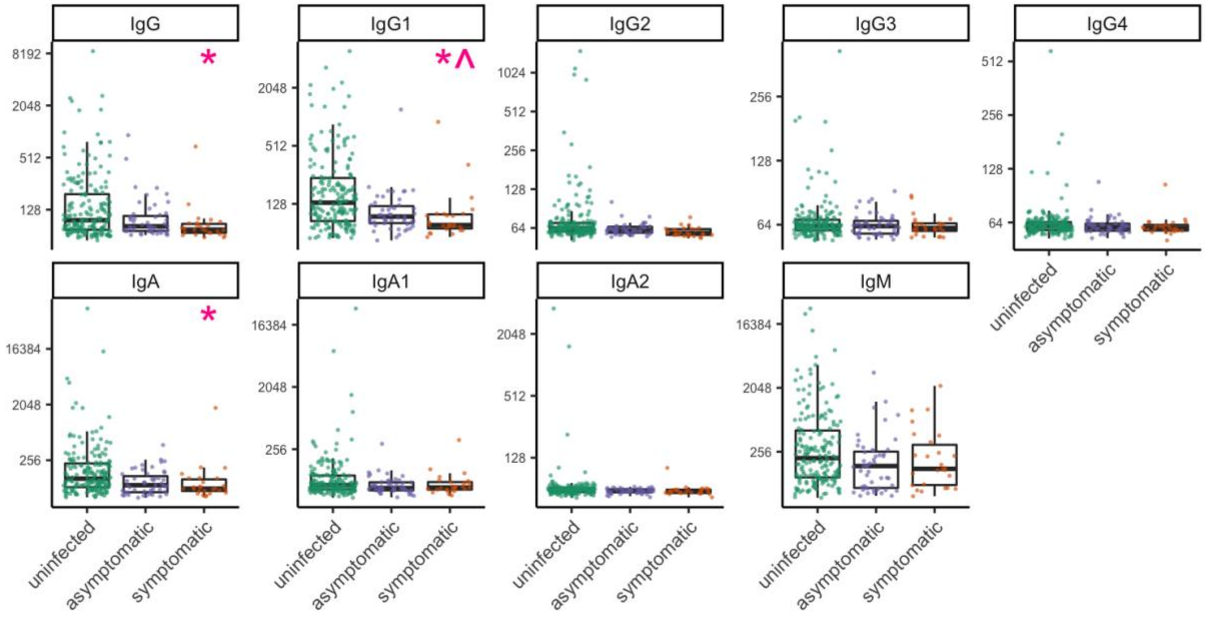


Inaba-OSP

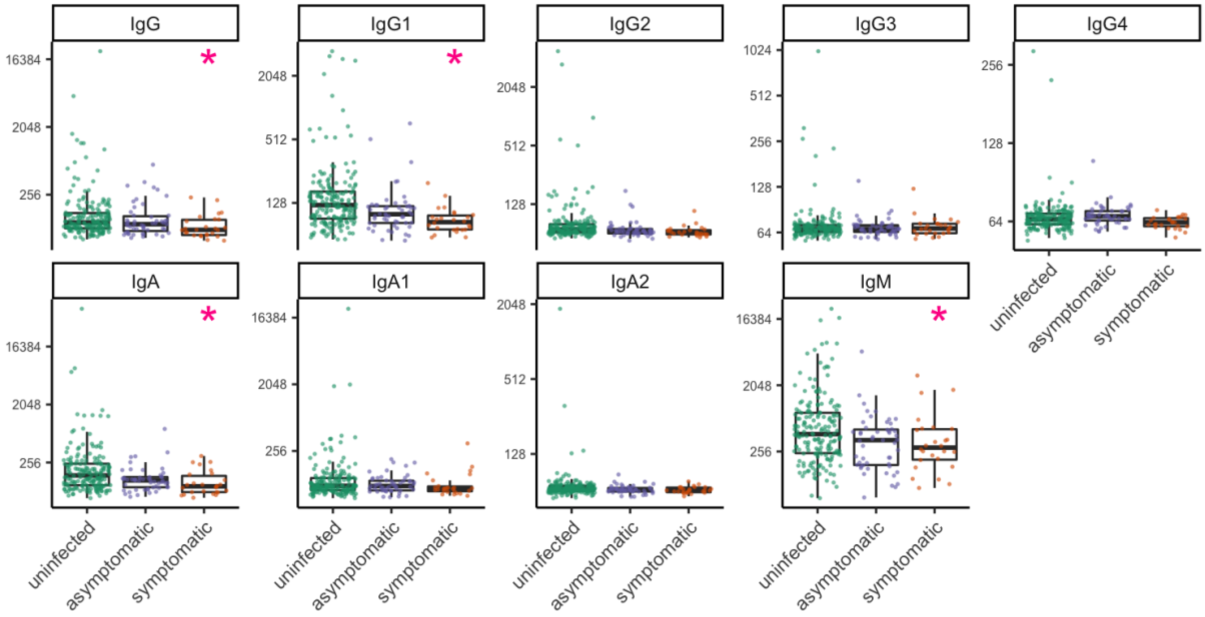

**Figure S2. Risk of developing symptoms among infected household contacts**

Odds ratio of symptoms among infected household contacts (n=77) for every 2-fold increase in baseline (Day 2) antibody titers, after adjusting for age and household clustering. Mean and 95% confidence interval are shown for each biomarker analyzed independently. Arrows indicate where upper confidence interval extends beyond the graph. Isotype is indicated on the left and antigen on the right for binding antibody titers. Biomarkers for which logistic model fitted values were very close to 1 are excluded.

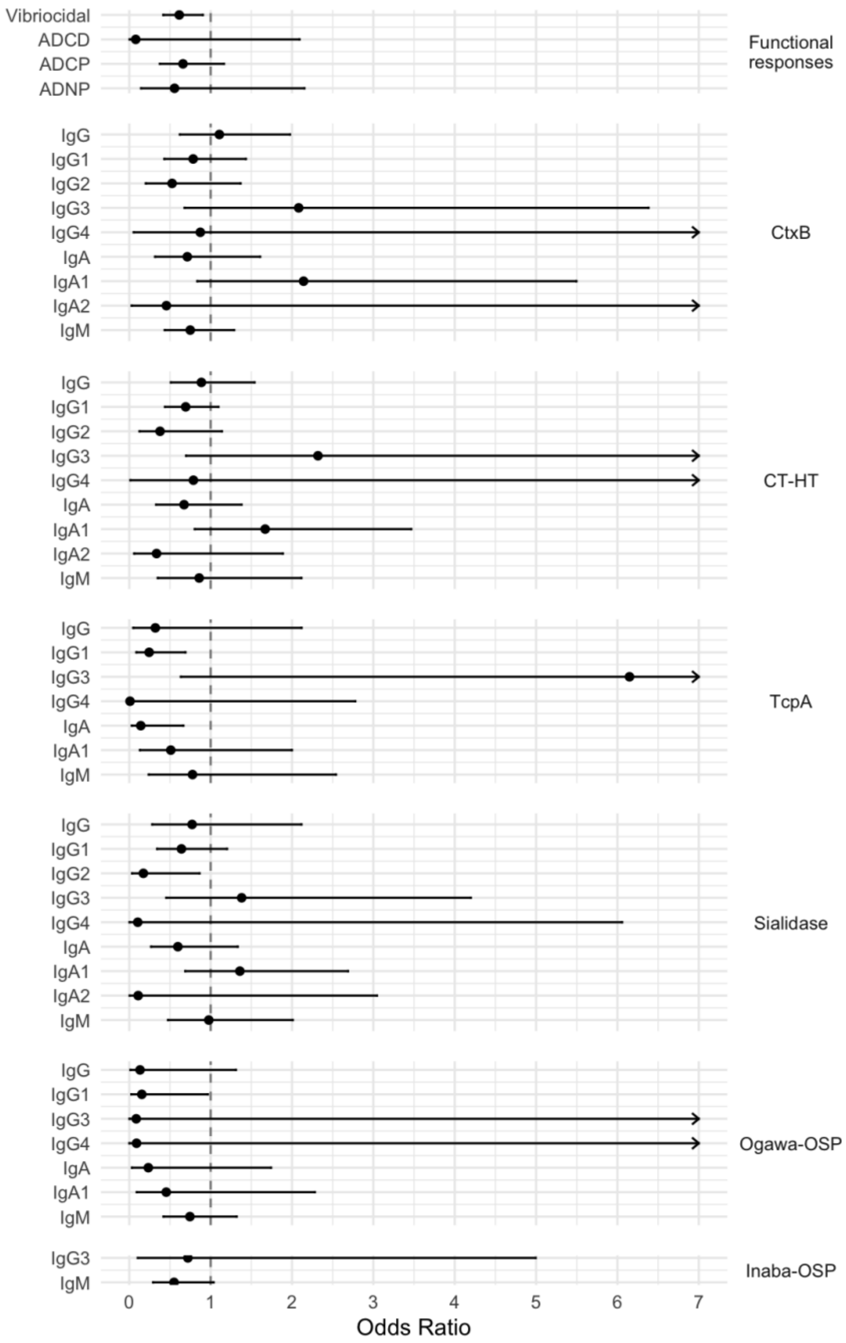

**Figure S3. Unsupervised analysis of biomarkers in household contacts of index cholera cases upon enrollment**

Generalized principal components analysis (GLM-PCA) plot for dimension reduction of the antibody responses in household contacts of index cholera cases upon enrollment (Day 2) (Townes et al. 2019). The first two components are shown, labelled by individual outcome.

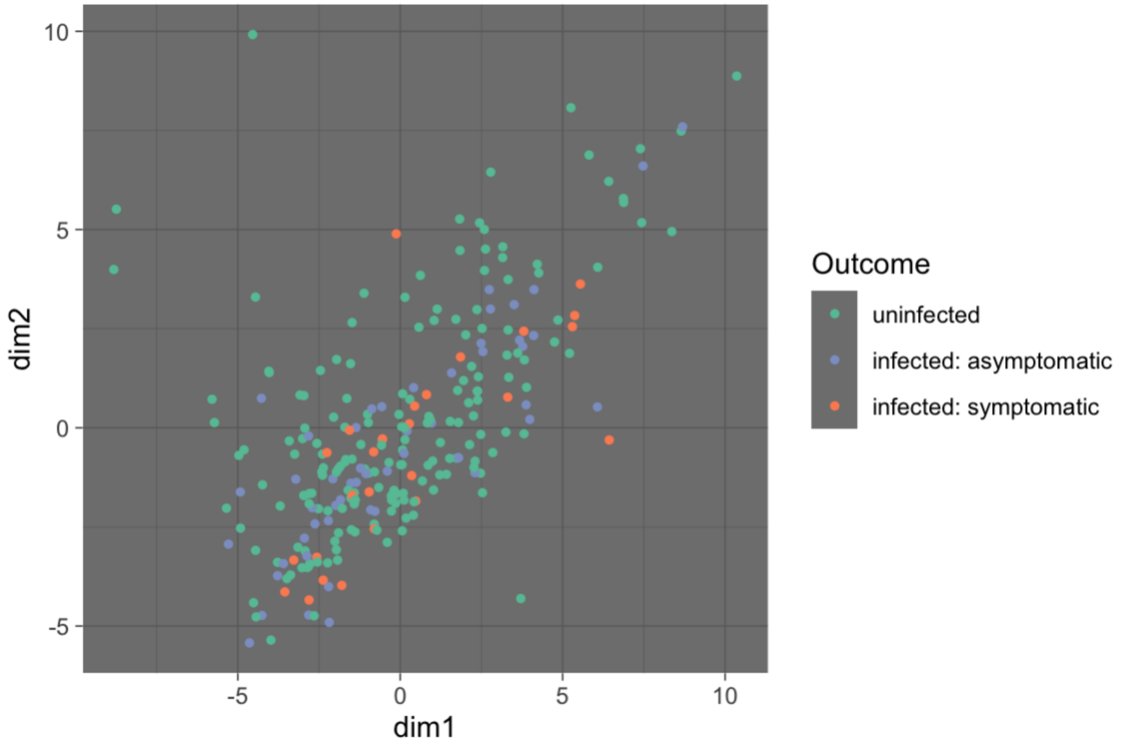

**Figure S4. Pair-wise correlations between biomarkers in household contacts**

Pair-wise Spearman rho coefficients for serum biomarkers among household contacts of cholera cases upon enrollment. Significant (p < 0.05) correlations are shown by an open box and non-significant correlations are marked with an X.

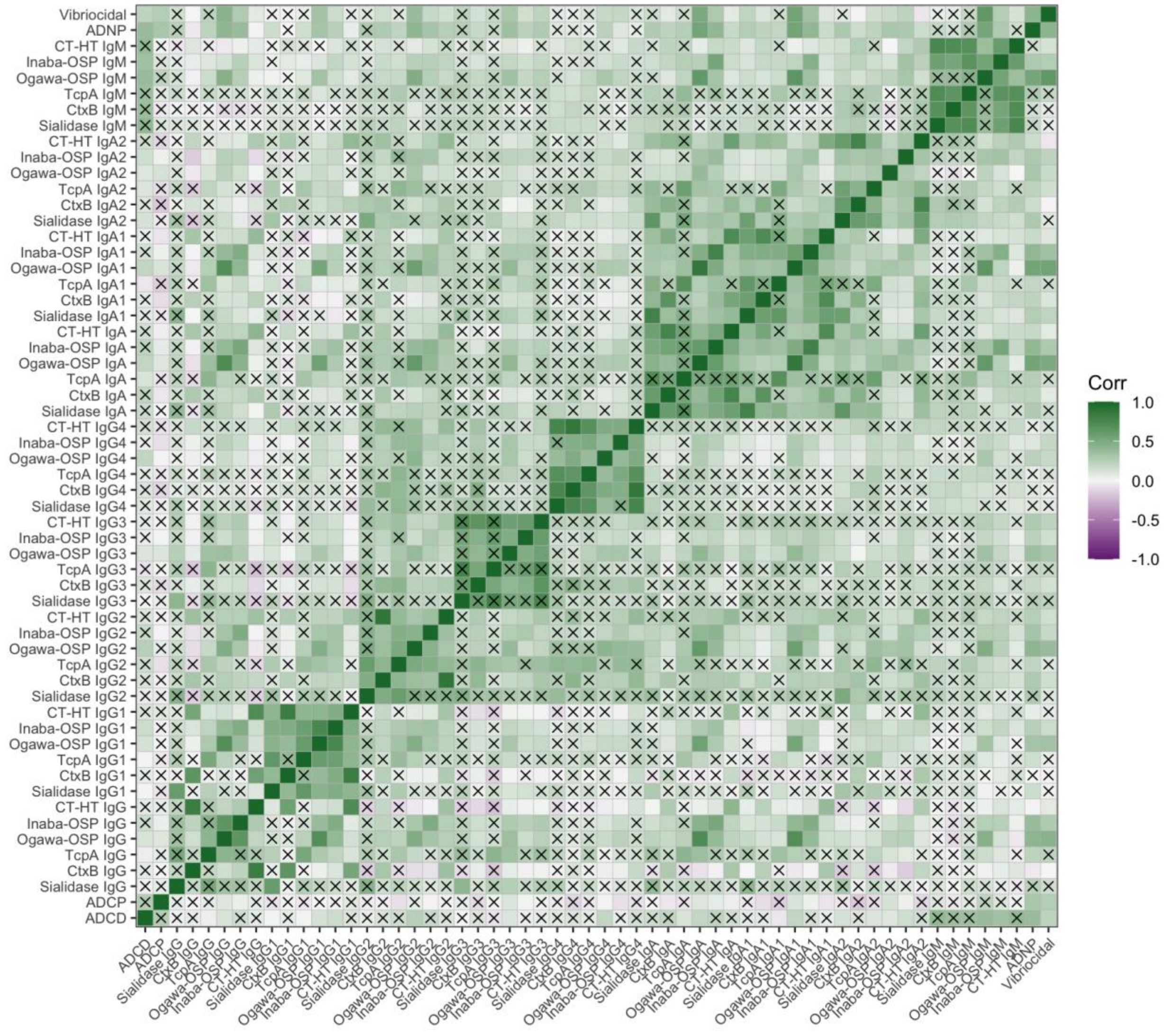

**Figure S5. Predicting whether household contacts remain uninfected using an ensemble of machine learning models**

**A)** Cross-validated receiver operator curves (cvROC) for classifying household contacts of index cholera cases that remain uninfected vs. become infected using an ensemble of three machine-learning models: random forest, penalized logistic regression, and support vector machine. Models were run with different subsets of biomarkers and age. “5 biomarkers” corresponds to five of the top biomarkers selected via conditional importance, including ADCD, CtxB IgM, CT-HT IgG1, Ogawa-OSP IgG1, TcpA IgG2. True and false positive rates calculated using leave-one-out cross-validation. **B)** Cross-validated area under the curve (cvAUC) corresponding to the models in A. Influence-curve based 95% confidence intervals are shown for the cvAUC estimates.

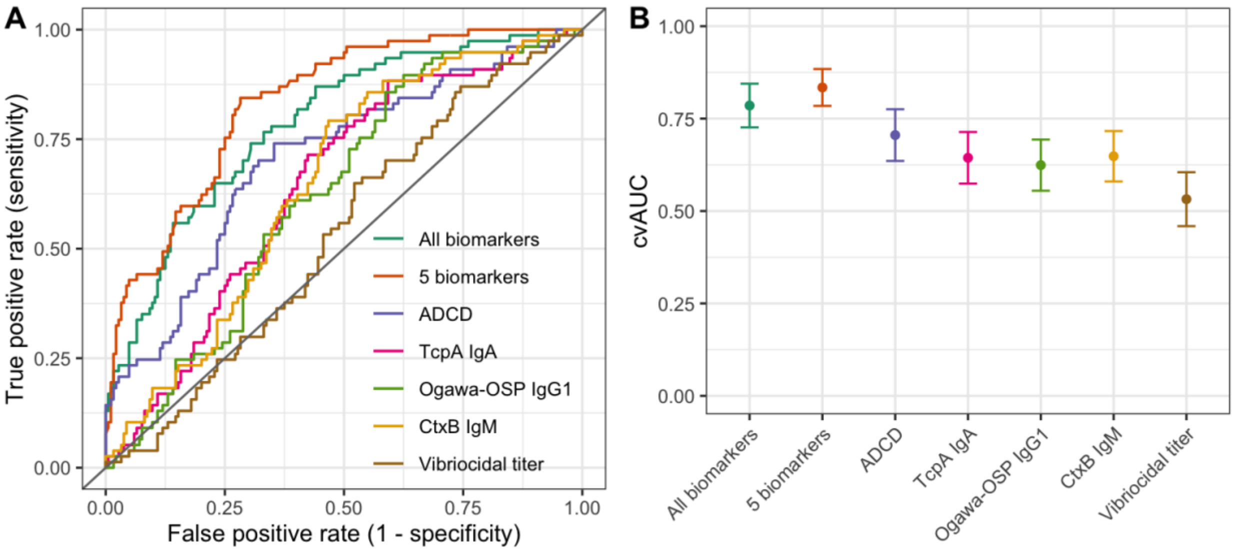

**Figure S6. Unsupervised analysis of biomarkers in vaccinees upon *V. cholerae* challenge**

Generalized principal components analysis (GLM-PCA) plot for dimension reduction of the antibody responses in volunteers that received a live attenuated oral cholera vaccine prior to challenge with *V. cholerae* (Townes et al. 2019). The first two components are shown, labelled by individual outcome. Results are shown for antibody responses on the day of challenge.

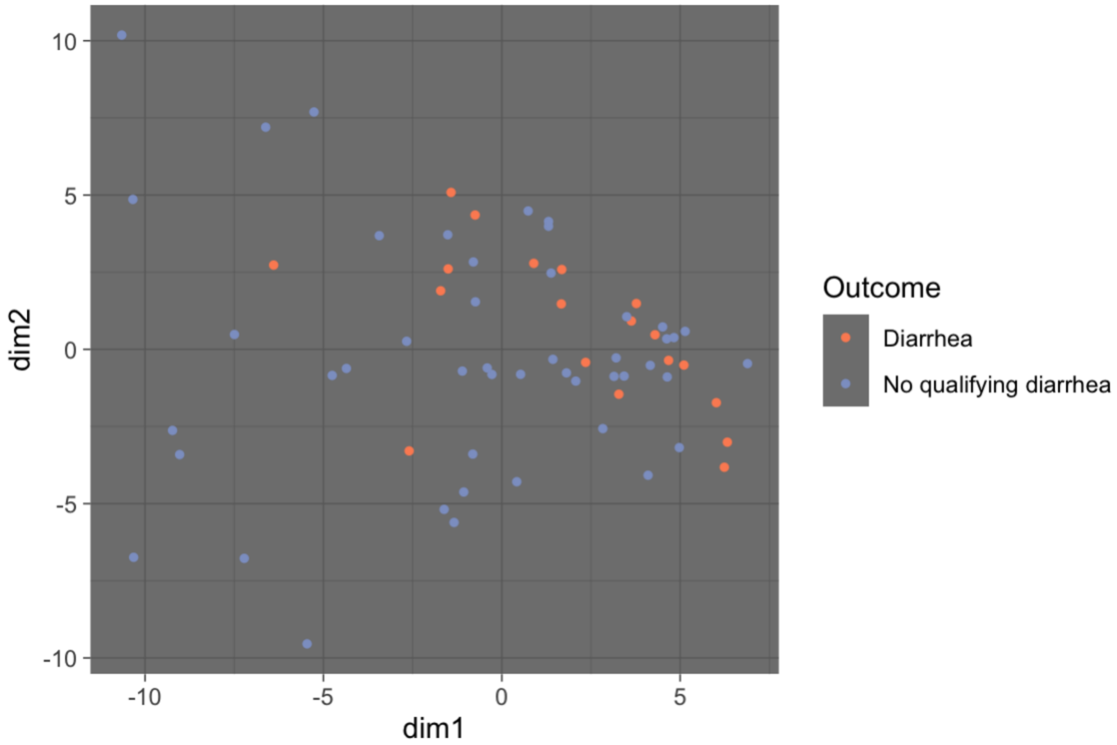

**Figure S7. Pair-wise correlations between biomarkers in vaccinees**

Pair-wise Spearman rho coefficients for serum biomarkers among vaccinees prior to *V. cholerae* challenge. Significant ($p < 0.05$) correlations are shown by an open box and non-significant correlations are marked with an X.

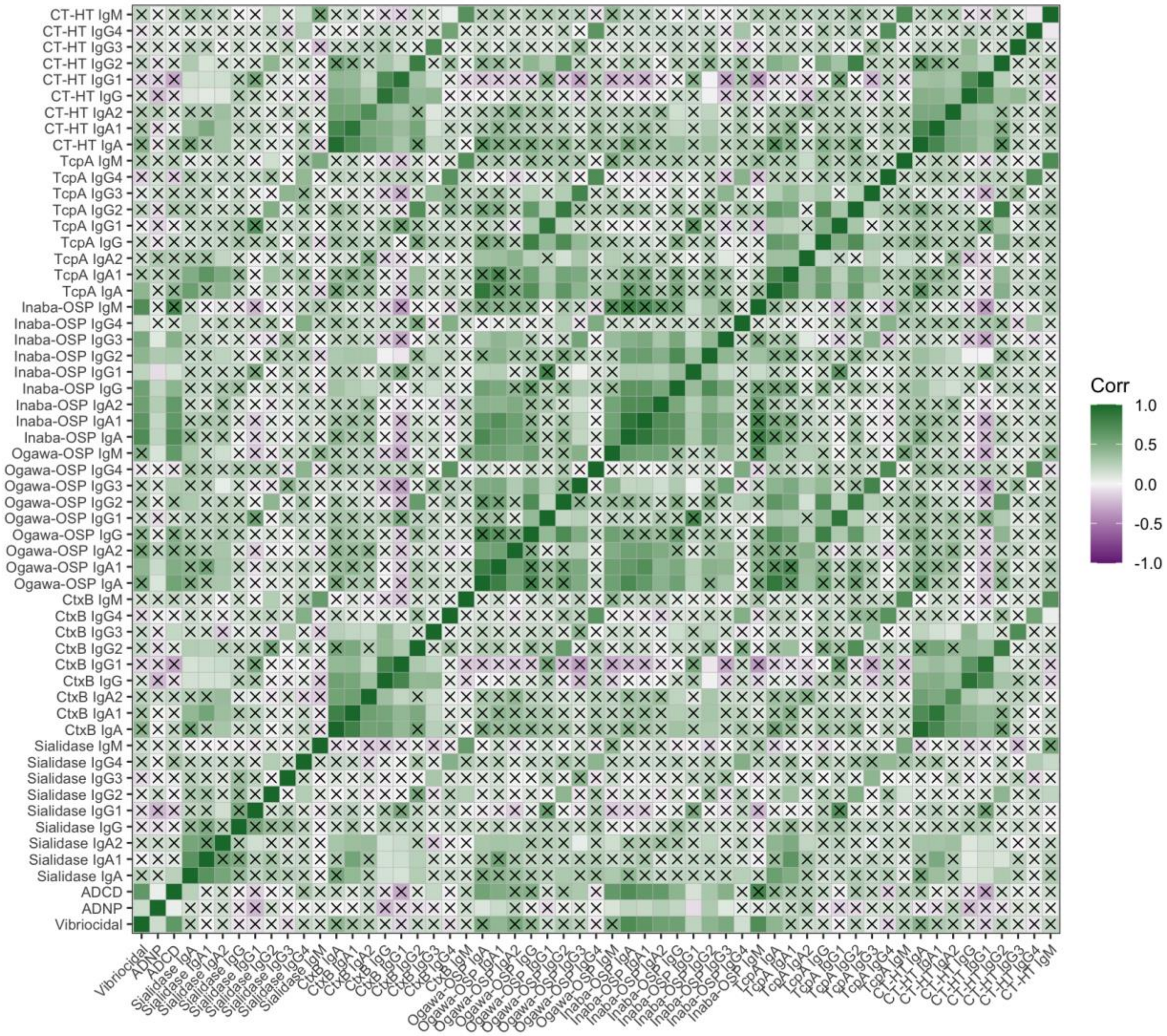

**Figure S8. Biomarkers important for classifying household contacts ages 18-48 by infection outcome**

Top 15 biomarkers important in classifying household contacts aged 18-48 years old as infected (i.e., becoming stool culture positive) vs. uninfected (i.e., remaining stool culture negative). Biomarkers are ranked by importance scores calculated using conditional random forest classification models.

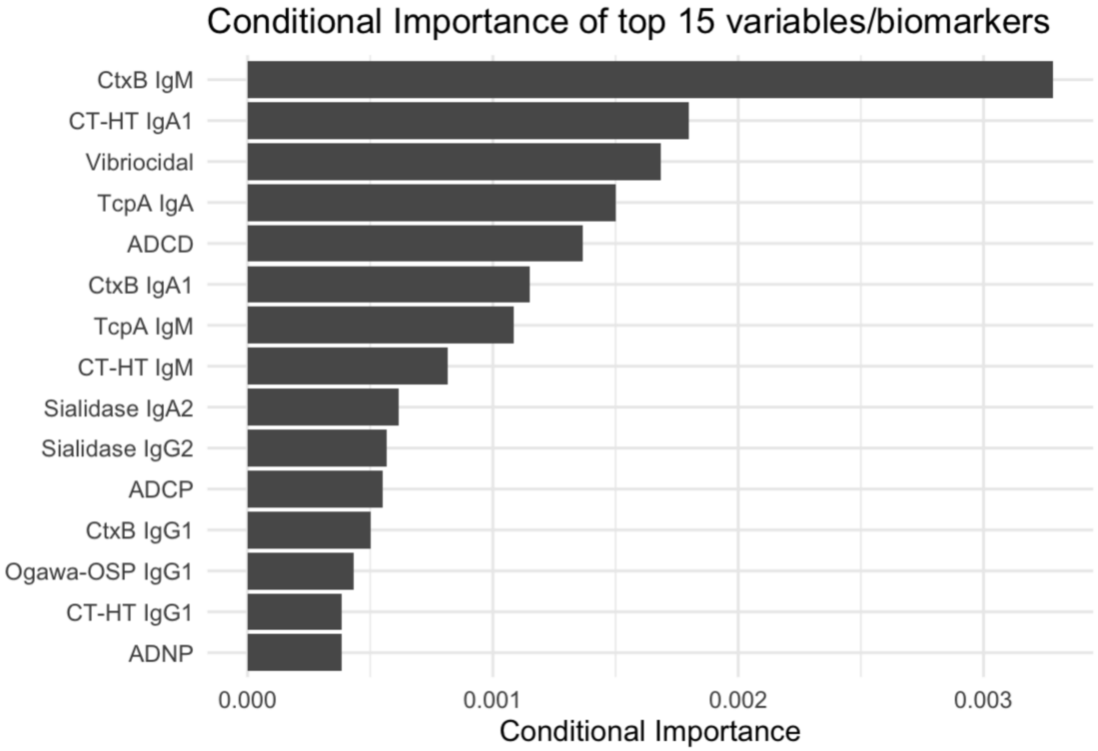

**Figure S9. Biomarkers important for classifying vaccinees by outcome following challenge on either day 10 or day 90**

Top 15 biomarkers important in classifying vaccinees by developing either no qualifying diarrhea or mild to severe diarrhea following *V. cholerae* challenge on day 10 **(A)** or day 90 **(B)**. Biomarkers were selected using serum collected from participants on the day of challenge. Biomarkers are ranked by importance scores calculated using conditional random forest classification models.

**A) Day 10**

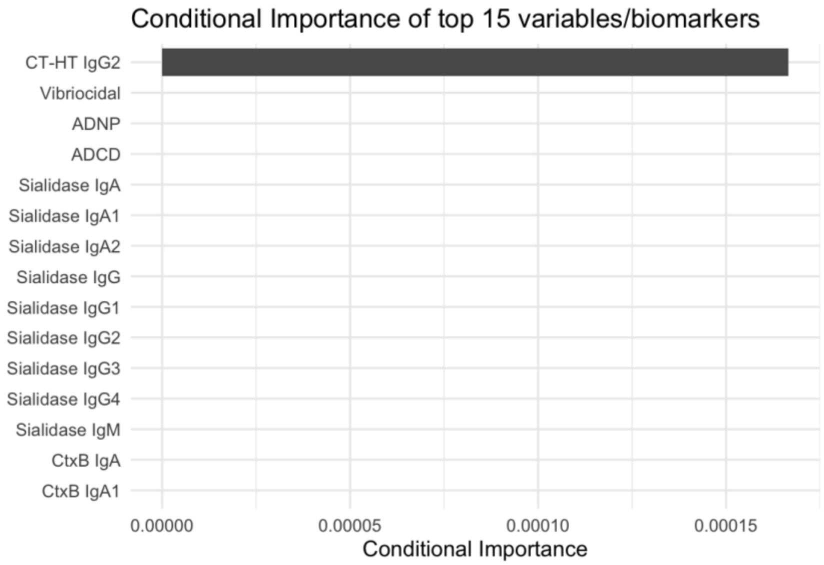


**B) Day 90**

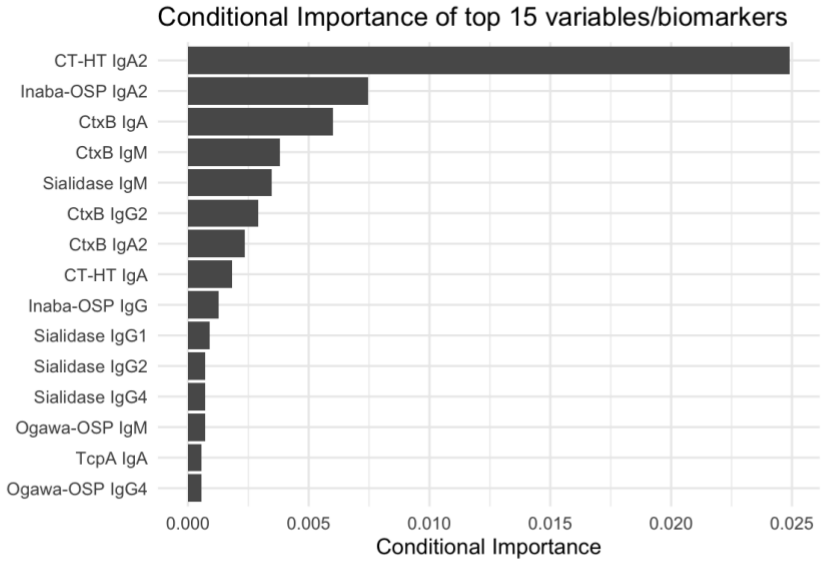

**Figure S10. Predicting whether vaccinees develop diarrhea following challenge on either day 10 or day 90**

**A)** Cross-validated receiver operator curves (cvROC) for classifying vaccinees that develop diarrhea vs. those that do not using random forest models with different subsets of biomarkers and age. “5 biomarkers” corresponds to five of the top biomarkers selected via conditional importance. For Day 10, “5 biomarkers” included vibriocidal titer, ADNP, ADCD, Sialidase IgA, and Sialidase IgA1. For Day 90, “5 biomarkers” included CT-HT IgA2, Inaba-OSP IgA2, CtxB IgA, CT-HT IgA, and CtxB IgM. True and false positive rates calculated using leave-one-out cross-validation. **B)** Cross-validated area under the curve (cvAUC) corresponding to the models in A. Influence-curve based 95% confidence intervals are shown for the cvAUC estimates.

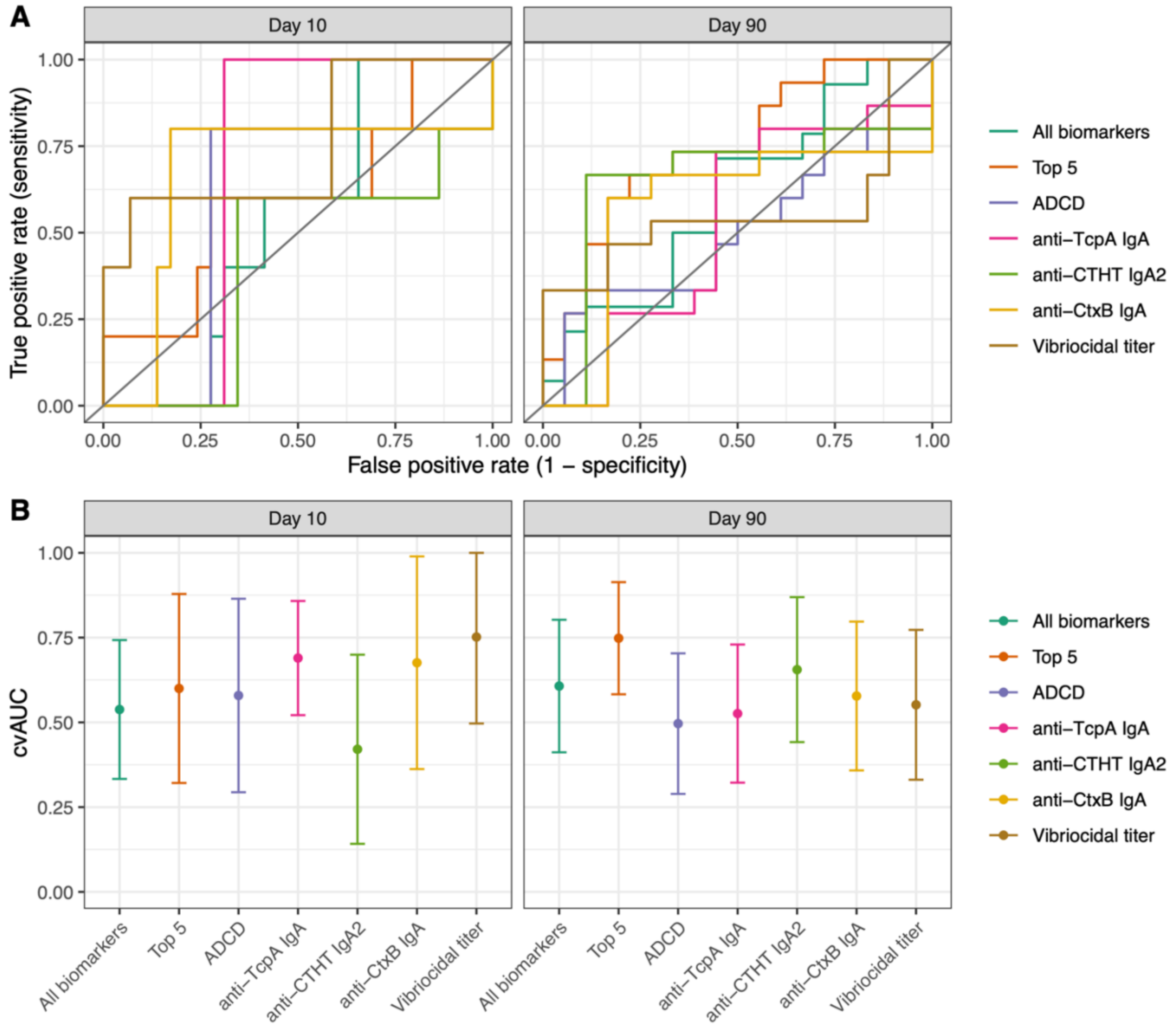

**Figure S11. Predicting whether vaccinees develop diarrhea using fold increase in vibriocidal titers**

**A)** Cross-validated receiver operator curves (cvROC) for classifying vaccinees that develop diarrhea vs. those that do not using conditional random forest models with different vibriocidal titer measurements and age. True and false positive rates calculated using leave-one-out cross-validation. **B)** Cross-validated area under the curve (cvAUC) corresponding to the models in A. Influence-curve based 95% confidence intervals are shown for the cvAUC estimates.

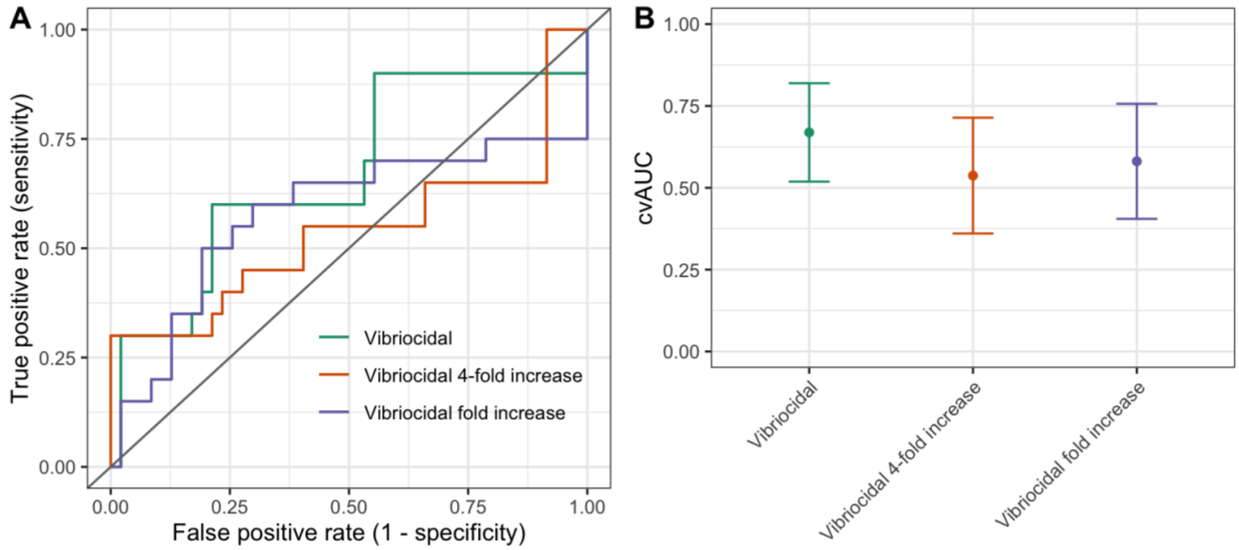

**Figure S12. Predicting whether vaccinees develop diarrhea using an ensemble of machine learning models**

**A)** Cross-validated receiver operator curves (cvROC) for classifying vaccinees that develop diarrhea vs. those that do not using an ensemble of three machine-learning models: random forest, penalized logistic regression, and support vector machine. Models were run with different subsets of biomarkers measured in serum on the day of challenge and age. “5 biomarkers” corresponds to five of the top biomarkers selected via conditional importance, including CT-HT IgA, CT-HT IgA2, CtxB IgA, TcpA IgA, and Inaba-OSP IgG3. True and false positive rates calculated using leave-one-out cross-validation. **B)** Cross-validated area under the curve (cvAUC) corresponding to the models in A. Influence-curve based 95% confidence intervals are shown for the cvAUC estimates.

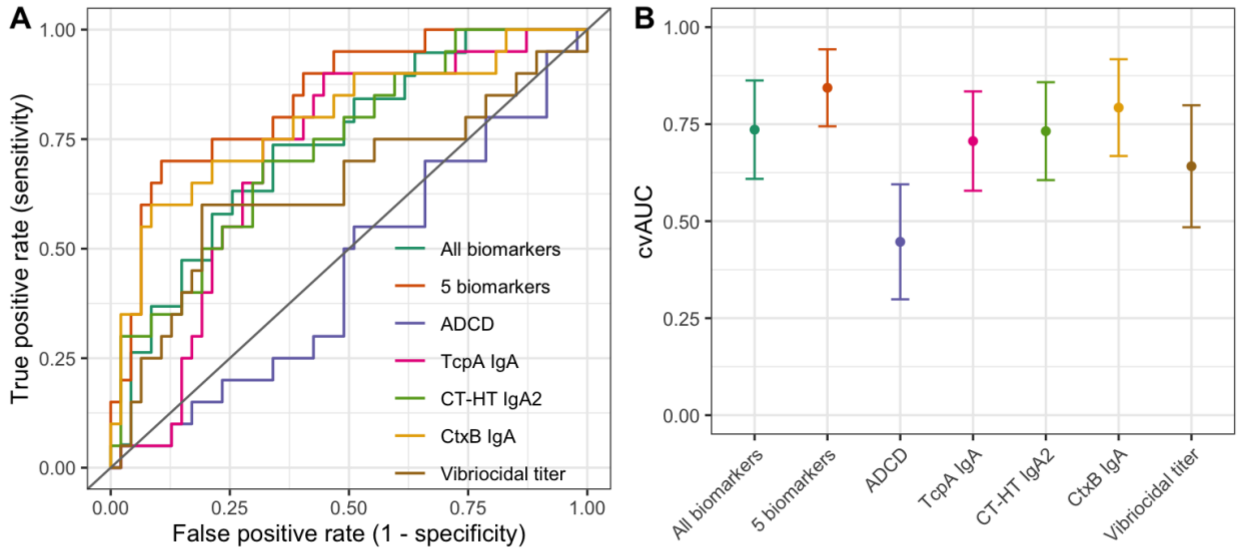

**Figure S13. Predicting infection outcome among household contacts using models fit with vaccinee data and vice versa**

**A)** Cross-prediction cross-validated receiver operator curves (cvROC). "Contacts predicted with vaccinees" indicates predicting whether household contacts remained uninfected vs. became infected using the "5 biomarkers" random forest model fit with the vaccinee data (biomarkers included ADCD, CtxB IgM, TcpA IgG2, Ogawa-OSP IgG1, and Sialidase IgG1). "Vaccinees predicted with contacts" indicates predicting whether or not vaccinees develop diarrhea using the "5 biomarkers" random forest model fit with the household contacts data (biomarkers included CT-HT IgA, CtxB IgA, CT-HT IgA2, TcpA IgA, and Sialidase IgA2). True and false positive rates calculated using leave-one-out cross-validation. **B)** Cross-validated area under the curve (cvAUC) corresponding to the models in A. Influence-curve based 95% confidence intervals are shown for the cvAUC estimates.

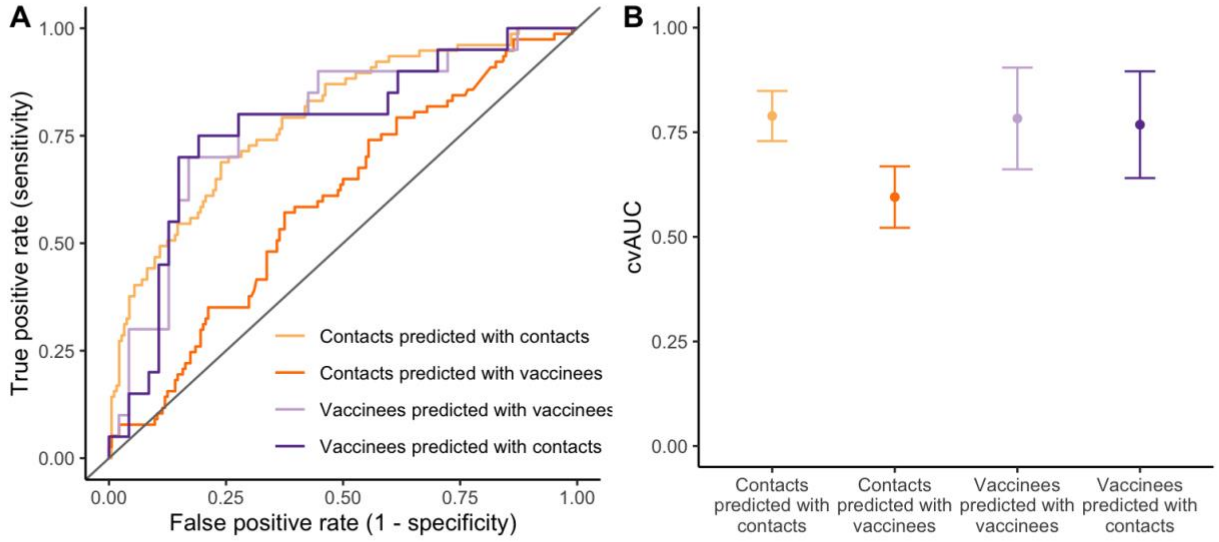

**Figure S14. Comparison of original vibriocidal titer responses to new vibriocidal titers in household contacts of index cholera cases**

Vibriocidal antibody titers for the subset (n = 252) of samples for which there was sufficient sample remaining to re-run the vibriocidal antibody assays. Purple line shows a 1:1 relationship and the size of each point corresponds to the number of samples that fell at each point on the graph.

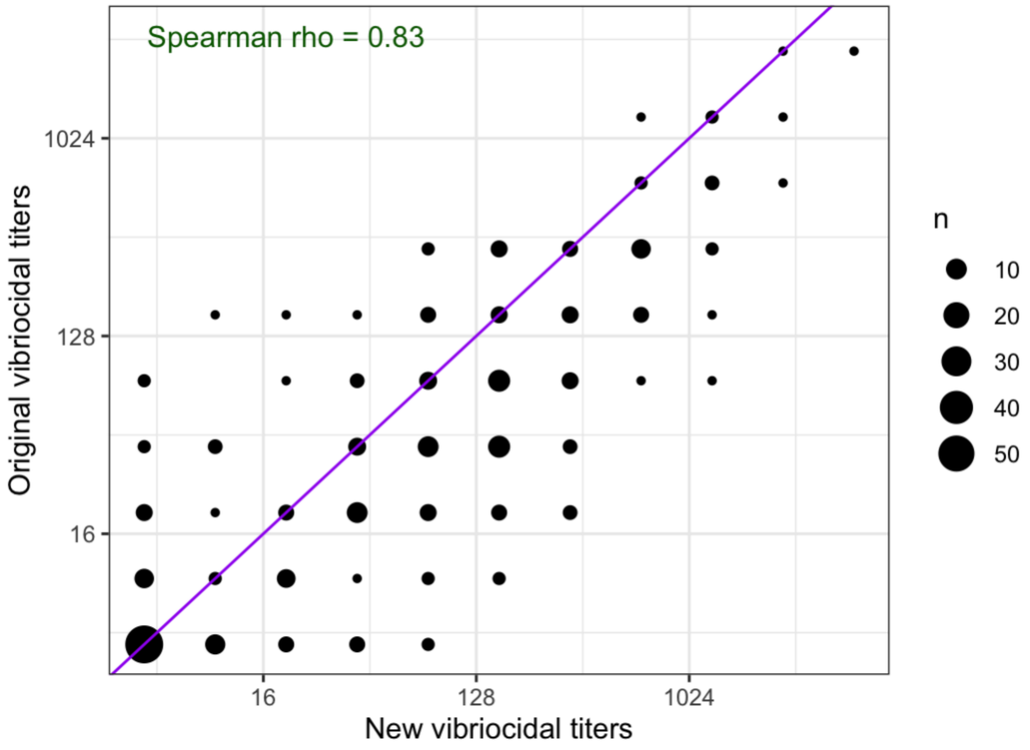

**Figure S15. Predicting stool culture status using old and new vibriocidal titers from the household contacts of index cholera cases**

**A)** Cross-validated receiver operator curves (cvROC) for classifying household contacts as uninfected vs. infected based on old and new vibriocidal antibody titers and age. Analysis includes the subset (n = 252) of samples for which there was sufficient sample remaining to re-run the vibriocidal antibody assays. True and false positive rates calcualted using leave-one-out cross-validation. **B)** Cross-validated area under the curve (cvAUC) corresponding to the models in A. Influence-curve based 95% confidence intervals are shown for the cvAUC estimates.

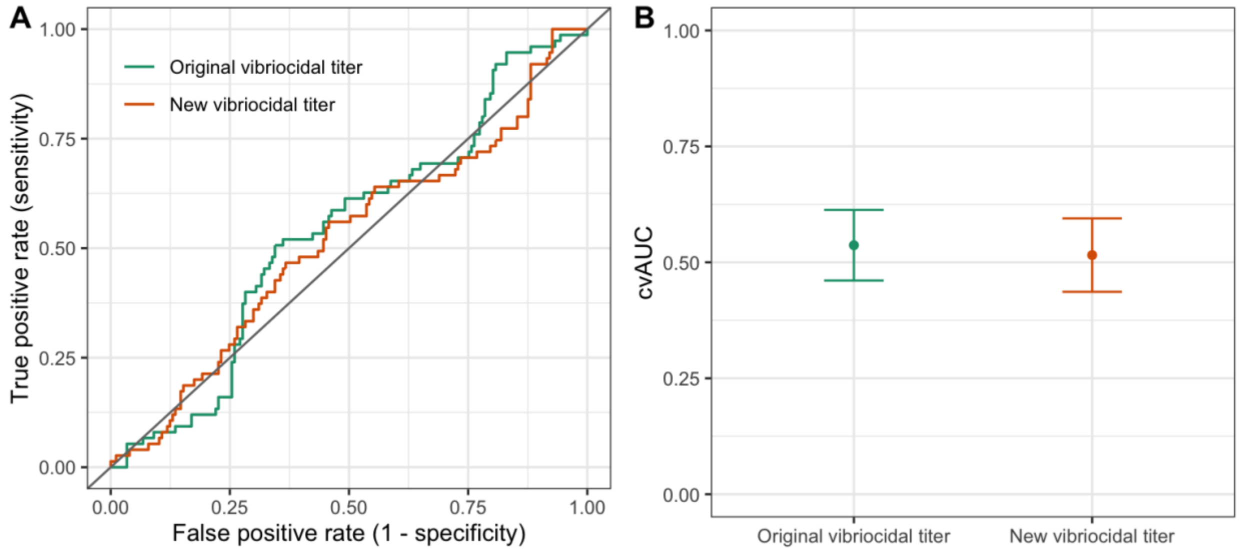
